# LIFE: A Deep Learning Framework for Laboratory Data Imputation in Electronic Health Records

**DOI:** 10.1101/2023.10.31.23297843

**Authors:** Samuel P. Heilbroner, Curtis Carter, David M. Vidmar, Erik T. Mueller, Martin C. Stumpe, Riccardo Miotto

## Abstract

Laboratory data in electronic health records (EHRs) is an effective source of information to characterize patient populations, inform accurate diagnostics and treatment decisions, and fuel research studies. However, despite their value, laboratory values are underutilized due to high levels of missingness. Existing imputation methods fall short, as they do not fully leverage patient clinical histories and are commonly not scalable to the large number of tests available in real-world data (RWD). To address these shortcomings, we present Laboratory Imputation Framework using EHRs (LIFE), a deep learning framework based on multi-head attention that is trained to impute any laboratory test value at any point in time in the patient’s journey using their complete EHRs. This architecture (1) eliminates the need to train a different model for each laboratory test by jointly modeling all laboratory data of interest; and (2) better clinically contextualizes the predictions by leveraging additional EHR variables, such as diagnosis, medications, and discrete laboratory results. We validate our framework using a large-scale, real-world dataset encompassing over 1 million oncology patients. Our results demonstrate that LIFE obtains superior or equivalent results compared to state-of-the-art baselines in 23 out of 25 evaluated laboratory tests and better enhances a downstream adverse event detection task in 7 out of 9 cases, showcasing its potential in efficiently estimating missing laboratory values and, consequently, in transforming the utilization of RWD in healthcare.

## Introduction

Electronic health records (EHRs) create novel opportunities to leverage clinical information across large populations using machine learning and other data-driven approaches.^1^ EHRs contain a mix of structured and unstructured data elements, such as demographics, clinical diagnoses, laboratory results, medication prescriptions, and free-text clinical notes. Among these, laboratory data is particularly informative since it captures an objective physiological attribute of patients, rather than being used primarily for billing purposes such as International Classification of Diseases (ICD) diagnosis codes.^2^

However, there are many challenges associated with using laboratory results in real-world data (RWD) analysis. Foremost amongst these challenges is data missingness. Laboratory values can be missing from a patient’s EHR for various reasons, including patients not needing specific tests, irregular check-ups or follow-ups, data recording and integration issues, and more, often resulting in biased statistics and suboptimal clinical model performances, specifically in groups with traditionally poor access to healthcare.^3–5^

The task of imputing missing laboratory values is particularly difficult in EHRs, which commonly contain irregular time intervals between consecutive records. The literature presents a number of approaches, generally using either simple rules-based methods, statistical modeling, or machine learning. Some common statistical methods in use are expectation maximization, regression, and multiple imputations by chained equations (MICE).^6^ Machine learning techniques include tree-based models, such as Multi-directional Approach for Missing Value Estimation in Multivariate Time Series Data (MD-MTS),^7^ Gaussian processes such as 3D-MICE,^8^ and deep learning architectures, such as Bidirectional Recurrent Imputation for Time Series (BRITS),^9^ Multi-directional Recurrent Neural Networks (M-RNN),^10^ and Gated Recurrent Unit with Decay.^11^ A recent benchmark suggested that MD-MTS may have the best performance on healthcare data.^12^

All these methods have limitations. First, they have been trained and evaluated on clean, homogenous, regularly sampled ICU data from a single institution, such as the Medical Information Mart for Intensive Care (MIMIC) datasets,^13,14^ and have not been tested on the heterogeneous, irregularly sampled, outpatient and inpatient RWD from multiple institutions, which is more common in the healthcare industry. Second, none of them take advantage of the entire feature space available in the EHRs. MICE is designed for cross-sectional data and is not well suited to longitudinal EHRs because it does not consider patient’s past and future laboratory values.^15^ Methods like BRITS, MD-MTS, and M-RNN consider past and future values of the laboratory test of interest, but ignore other EHR variables, such as demographics, ICD codes, and discrete laboratory results (e.g., EGFR status), which can be useful to clinically contextualize the imputed values. Last, these methods were designed to either infer one laboratory value at the time or, because of architectural constraints, a small subset of values (up to 25-50 different tests). However, there are about 31,000 different laboratory tests in the Logical Observation Identifiers Names and Codes (LOINC) catalog and building one model for each of them is impractical.

Recent advances in deep learning^16^ and self-supervised learning^17^ are enabling new ways to leverage EHRs for personalized healthcare,^18–20^ and provide an opportunity to address the challenges discussed above. The transformer architecture and its building block, the multi-head self-attention module, specifically, can contextualize each data point more efficiently than other machine learning architectures^21^, showing improved performances when modeling EHRs.^22^ This work fits into this landscape by presenting a scalable architecture based on self-supervised learning to process heterogeneous and large cohorts of clinical data for imputing laboratory data into patient clinical histories.

In particular, this paper proposes Laboratory Imputation Framework using EHRs (LIFE), a novel and scalable architecture that can impute any laboratory value at any point in time from EHRs (Figure 1). LIFE receives as input a patient’s EHR data and a query describing the laboratory test of interest. The model leverages multi-head attention and time decay layers and a training objective inspired by masked language modeling (Figure 2). The framework uses the entire feature space of the EHRs, including discrete laboratory values and codes. We demonstrate that LIFE performs better than several baselines including MICE, BRITS, and MD-MTS using real-world multi-institute EHRs of about 1.1 million de-identified oncology patients in the Tempus data warehouse. We then show that this improvement in imputation performance translates to better performance on a downstream task, namely detection of adverse events. Finally, using attention weights, we show that LIFE is interpretable and the feature importances it selects are consistent with medical knowledge.

**Figure 1:**
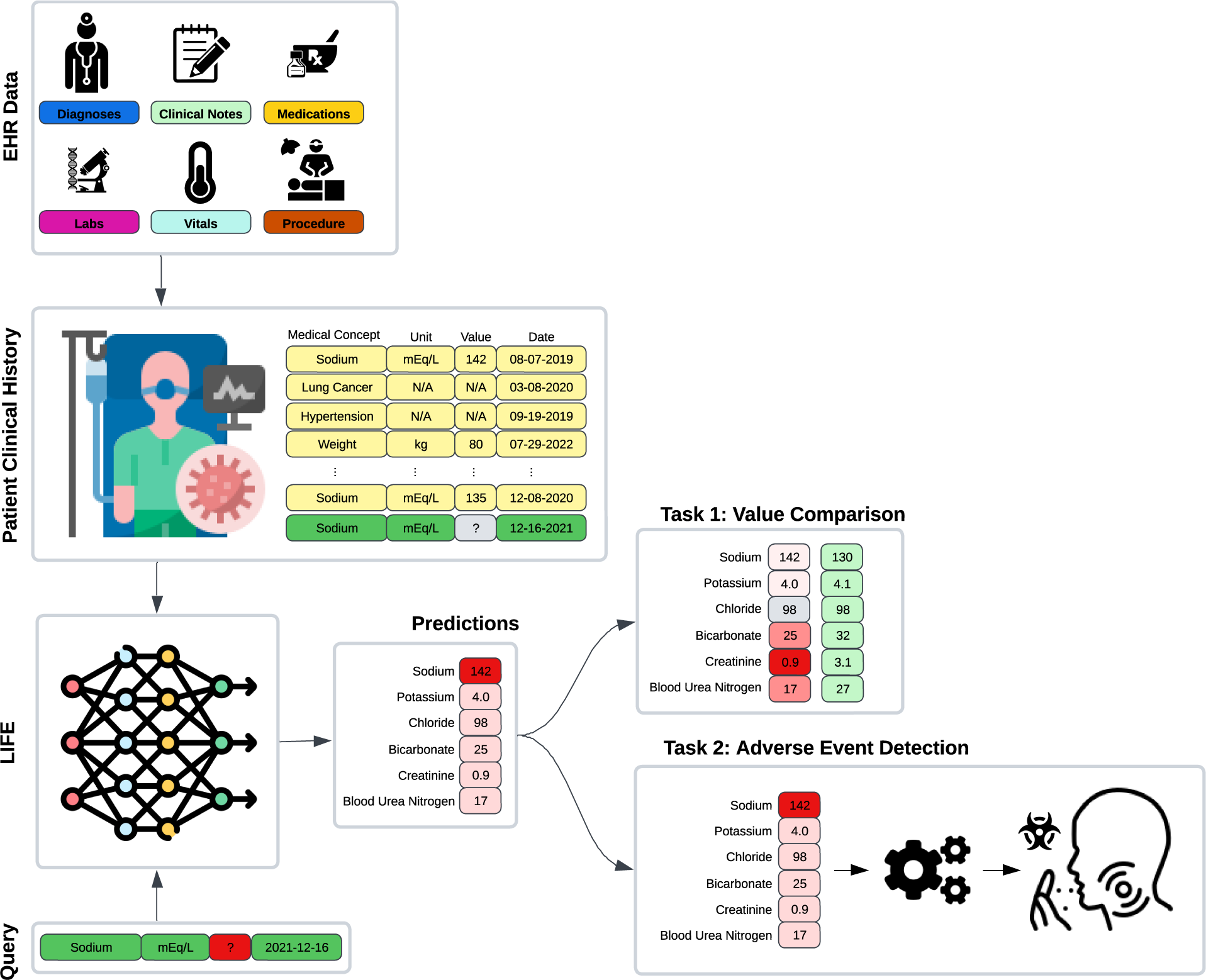
Overview of the LIFE experimental framework. EHR observations and a query comprising a laboratory test, a unit of measure, and a date are input into LIFE to impute the corresponding laboratory value for that specific time point. We assessed these imputations within two scenarios. For the initial experiment (Task 1), we masked laboratory data from a patient’s clinical history, replaced the absent value through imputation, and then compared it with the actual measurement. In the subsequent experiment (Task 2), we employed LIFE’s laboratory value imputations as features within a downstream task focused on detecting adverse events.

**Figure 2:**
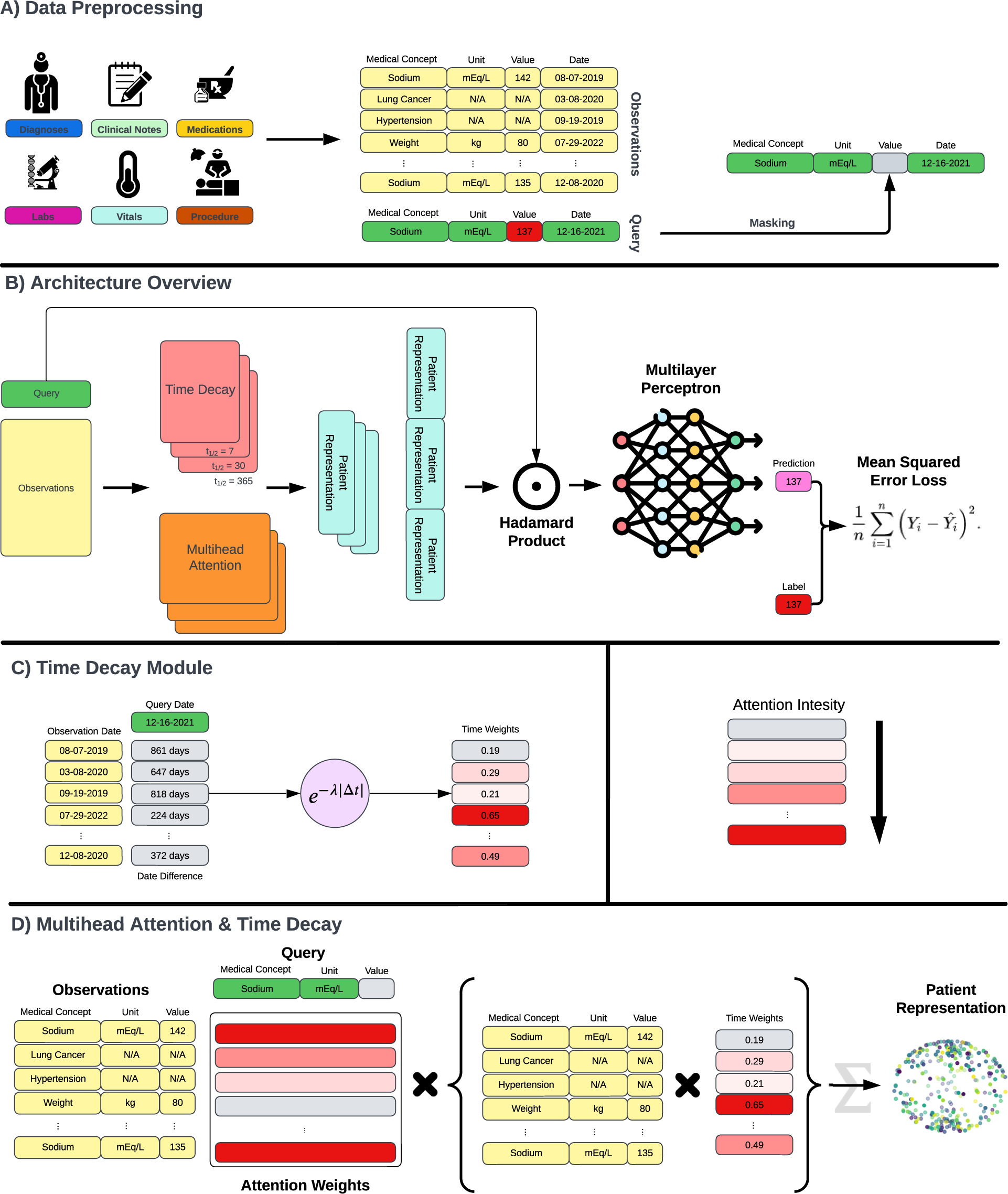
LIFE Model Architecture. (A) The model’s inputs consist of a query, which includes a laboratory test, a unit of measure, and a date, along with EHR observations extracted from patient clinical records. During the training phase, each query is chosen randomly from the available patient laboratory data, and its value is masked before being fed into the model. (B) LIFE is constructed with time decay and multi-head attention layers, both contributing to the creation of patient embeddings. The multiple time decay modules compute observations across various time scales. The patient embeddings resulting from this process are then subjected to multiplication by the query using the Hadamard product. The concatenated result is passed through a multi-layer perceptron to generate a final prediction. Throughout the training iterations, the model predicts the masked values and aims to minimize the discrepancy between original and imputed values through Mean Square Loss. (C) The time decay module assigns weights to observations based on their proximity to the time point specified in the query. These weights are determined by calculating the time difference in days between each observation and the query and then applying an exponential decay function. (D) The multi-head attention module assigns weights to observations according to their medical significance relative to the query. This module takes three inputs: the query, the observations, and the weights computed by the time decay mechanism.

This architecture enables laboratory data imputation at scale by eliminating the need for training a different model for each laboratory test and by including the use of additional EHR variables to better clinically contextualize the predictions. By imputing a large amount of laboratory data for patients at different points in time, LIFE aims to reduce the sparseness of real-world clinical data and provide a more complete and context-aware characterization of patient clinical histories. This will consequently enhance systems using EHRs for, among others, clinical decision support, care gaps detection, phenotyping, and clinical trial matching.^23–26^

## Results

We first assessed the capability of LIFE to impute laboratory data at scale. Specifically, we investigated the model’s ability to accurately infer randomly deleted laboratory values within patients’ clinical histories. To achieve this, we evaluated its performance on a hold-out test set of 110,953 patients across 344 laboratory tests, each of which with at least 10,000 occurrences in the training data (refer to the “Dataset” subsection in “Methods” for more details). Figure 3 and Supplementary Table 1 present results in terms of mean absolute error (MAE) for all tests. Overall, LIFE obtained strong performances, with an average MAE of 0.15. These results showcase LIFE’s ability to make effective predictions at scale. It is important to notice that these results were obtained using a single model that was jointly trained via masking to predict values for the 344 laboratory tests.

**Figure 3.**
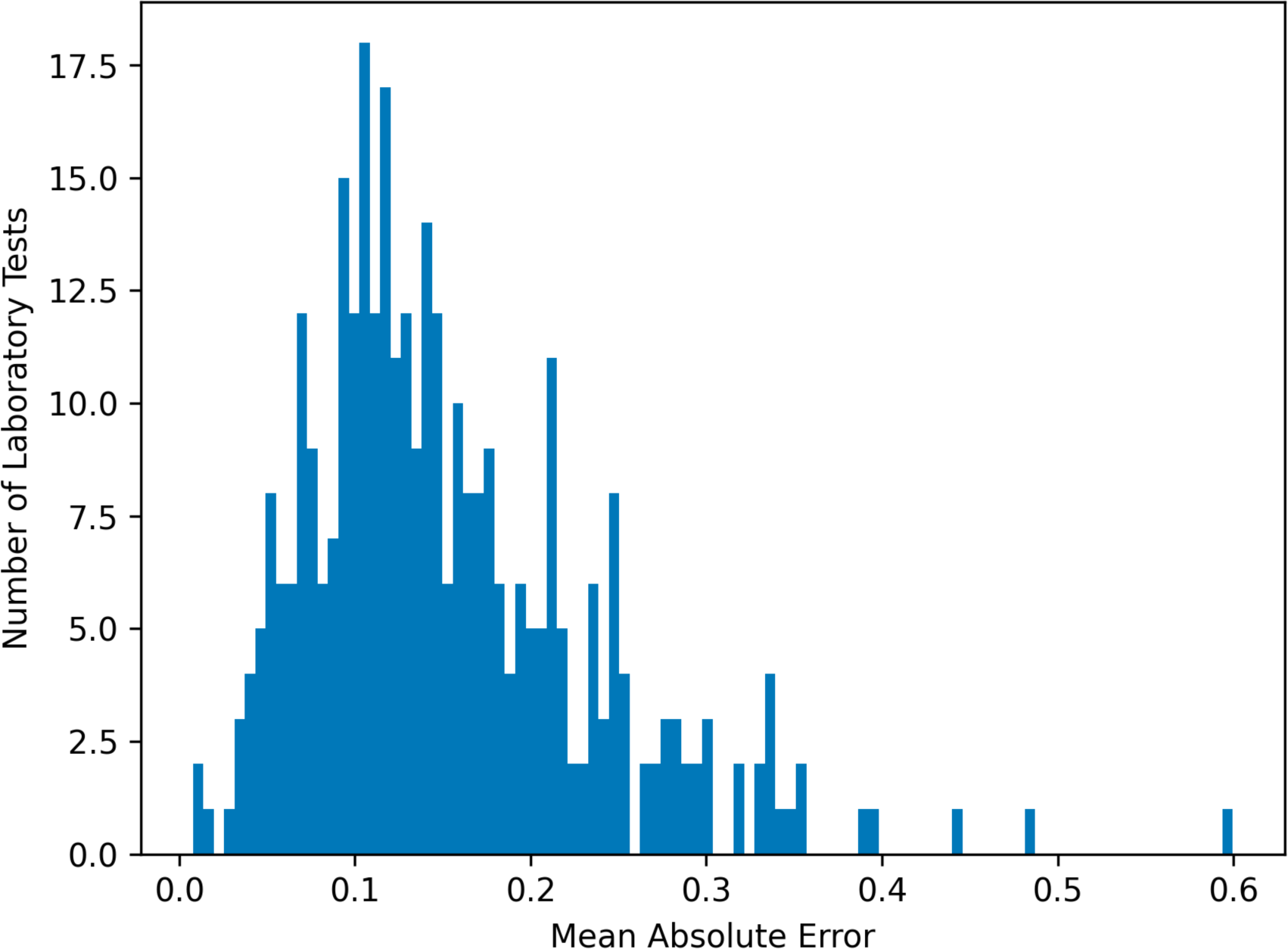
Performance of LIFE across 344 laboratory tests. The x-axis represents mean absolute error (MAE), and the y-axis displays the count of tests where LIFE reached the corresponding performance level. As evident from the graph, LIFE consistently performs effectively across all laboratory tests, implying its proficiency in imputing not only common tests but also a broader range including less frequent tests. The average MAE across all 344 tests was 0.15. All laboratory tests we standardized to a uniform distribution on [0, 1] to facilitate comparison.

Table 1 compares the performances of LIFE and several baselines from the literature on 25 common laboratory tests. Differently from LIFE, which by design can scale to any number of laboratory tests, baselines were only trained on these 25 common laboratory values. This corresponds to the approximate number of tests used to train and validate these models in the literature (see “Baselines” and “Experimental Design” subsections in “Methods” for more details). As it can be seen, LIFE outperformed all baselines with an average MAE of 0.14, while the second-best model, BRITS, had an average MAE of 0.16. LIFE also obtained the best performances in 23 out of the 25 laboratory tests and, notably, outperformed rules-based baselines by a substantial 19%.

**Table 1.** Laboratory data imputation performances of LIFE’s and baselines. LIFE achieved superior or equivalent results compared to the baselines in 23 out of 25 laboratory tests, assessed in terms of mean absolute error (MAE). This subset of laboratory data includes tests from the basic metabolic panel, complete blood count, vital signs, and other medically relevant tests, such as brain natriuretic peptide (BNP), troponin, hemoglobin A1c (HbA1c), and prostate-specific antigen (PSA). AST = aspartate aminotransferase; ALT = alanine transaminase; ALP = alkaline phosphatase. The first row is the average performance across all laboratory tests reported in the table.

| Laboratory Test | Interpolation | Nearest Lab | BRITS | MD-MTS | MICE | LIFE |
| --- | --- | --- | --- | --- | --- | --- |
| <u>Average</u> | 0.16 | 0.17 | 0.16 | 0.18 | 0.22 | **0.14** |
| Sodium | 0.19 | 0.20 | 0.15 | **0.14** | 0.19 | 0.15 |
| Potassium | 0.21 | 0.22 | 0.18 | 0.23 | 0.23 | **0.19** |
| Chloride | 0.18 | 0.18 | 0.14 | 0.15 | 0.19 | **0.13** |
| Bicarbonate | 0.20 | 0.20 | 0.24 | 0.18 | 0.24 | **0.15** |
| Urea Nitrogen | 0.16 | 0.17 | 0.14 | 0.17 | 0.20 | **0.13** |
| Creatinine | 0.12 | 0.12 | 0.12 | 0.13 | 0.20 | **0.10** |
| Hemoglobin | 0.13 | 0.13 | 0.11 | 0.14 | 0.22 | **0.06** |
| White Blood Cell | 0.16 | 0.16 | 0.15 | 0.17 | 0.21 | **0.09** |
| Platelet Count | 0.15 | 0.15 | 0.14 | 0.16 | 0.21 | **0.13** |
| Glucose | 0.20 | 0.21 | **0.18** | 0.20 | 0.23 | **0.18** |
| Calcium | 0.19 | 0.19 | **0.16** | 0.18 | 0.20 | **0.16** |
| AST | 0.17 | 0.17 | 0.16 | 0.16 | 0.17 | **0.13** |
| ALT | 0.16 | 0.16 | **0.13** | 0.16 | 0.18 | **0.13** |
| ALP | 0.13 | 0.14 | 0.14 | 0.16 | 0.22 | **0.12** |
| Albumin | 0.16 | 0.16 | 0.13 | 0.15 | 0.19 | **0.10** |
| Bilirubin | 0.16 | 0.17 | 0.16 | 0.18 | 0.23 | **0.15** |
| HbA1c | 0.22 | 0.22 | 0.21 | 0.27 | 0.21 | **0.16** |
| Troponin | 0.17 | 0.17 | 0.22 | 0.23 | 0.24 | **0.16** |
| BNP | 0.21 | 0.21 | 0.29 | 0.26 | 0.25 | **0.19** |
| PSA | 0.10 | **0.11** | 0.17 | 0.18 | 0.21 | **0.11** |
| Heart Rate | 0.18 | 0.19 | 0.18 | 0.18 | 0.24 | **0.17** |
| Respiration Rate | 0.21 | 0.21 | **0.18** | 0.22 | 0.24 | 0.20 |
| Oxygen | 0.21 | 0.22 | 0.21 | 0.22 | 0.24 | **0.20** |
| Height | **0.05** | **0.05** | 0.08 | 0.09 | 0.22 | **0.05** |
| Weight | 0.06 | 0.06 | 0.07 | 0.07 | 0.23 | **0.05** |

We notice that performances of LIFE remained consistent irrespective of the timing of prediction within a patient’s history (Supplementary Figure 1) and clinical status, i.e., tumor’s origin and stage (Supplementary Table 2). However, we observed a substantial variation in performance related to the magnitude of the laboratory value (Supplementary Figure 2). Specifically, LIFE exhibited better performances when predicting either very low or very high values for laboratory tests. This is actually to be expected: when the model is uncertain of its prediction, it is more likely to revert to the mean value of the test result. This hypothesis is supported by Supplementary Figure 3, which shows that predictions are biased towards central or median value for all models.

We also observe that LIFE’s continuous predictions can effectively categorize imputed values as normal or abnormal by assessing the extremeness of the predicted values. As illustrated in Supplementary Figure 4, LIFE outperformed all baselines in identifying abnormal laboratory values, achieving an area under the receiver operating characteristic curve (AUC-ROC) of 0.79 and an area under the precision - recall curve (AUC-PR) of 0.45.

We then evaluated the benefits of using LIFE’s imputed values in the downstream task of detecting adverse events from laboratory data. Specifically, for each patient, we imputed their laboratory data, which were then used to detect the presence of an adverse event at that time. As can be seen in Table 2, LIFE obtained the best AUC-PR for 7 out of 9 adverse events. On average, LIFE achieved an AUC-PR of 0.72, with the second best model for this use case, Nearest Lab Imputation, achieving 0.68 (6% improvement).

**Table 2.** LIFE and baseline performances on detecting adverse events. Laboratory values imputed by LIFE yielded superior performance in the downstream task of detecting adverse events compared to the baselines, as measured by the area under the precision-recall curve (AUC-PR). LIFE achieved an average AUC-PR of 0.72, while the second-best model, Nearest Lab Imputation, achieved 0.68 (a 6% improvement).

| Adverse Event | Interpolation | Nearest Lab | BRITS | MD-MTS | MICE | LIFE |
| --- | --- | --- | --- | --- | --- | --- |
| Sepsis | 0.67 | 0.68 | 0.66 | 0.67 | 0.67 | **0.70** |
| Anemia | 0.68 | 0.70 | 0.69 | 0.71 | 0.71 | **0.72** |
| Renal Failure | 0.69 | 0.71 | 0.70 | 0.69 | 0.69 | **0.72** |
| Transaminitis | 0.70 | 0.71 | 0.70 | 0.71 | 0.71 | **0.74** |
| Seizure | 0.67 | 0.68 | 0.67 | **0.75** | 0.75 | 0.73 |
| Febrile Neutropenia | 0.63 | 0.64 | **0.66** | 0.62 | 0.62 | **0.66** |
| Dehydration | 0.68 | **0.70** | 0.68 | 0.66 | 0.66 | 0.68 |
| Lung Infection | 0.65 | 0.66 | 0.66 | 0.66 | 0.66 | **0.71** |
| Cytopenia | 0.71 | 0.71 | 0.71 | 0.70 | 0.70 | **0.75** |

Lastly, we conducted an analysis of the attention weights assigned by LIFE to clinical features in the input space. Specifically, we looked at the attention weights assigned to six patients from the test set for different laboratory tests. For each patient, Figure 4 shows the 20 observations with the largest attention weights, accompanied by the corresponding laboratory test. Overall, we found that the attention weights assigned by the model were highly consistent with known medical knowledge, not limited to other laboratory data, which supports the validity of our model’s predictions. For example, as it can be seen, in the first patient who was queried for body weight, the model paid attention to past and future values of body weight, body surface area, and body mass index. In the case of hemoglobin, the model attended to hematocrit, which is closely related to hemoglobin, and mean corpuscular hemoglobin, which is often used to evaluate for anemia. Similarly, for leukocytes, the model attended to closely related laboratory tests in the white blood cell panel, such as the monocyte count. For albumin, the model attended to past and future albumin levels, as well as to total protein levels. Finally, in the case of prostate-specific antigen (PSA), the model attended to past and future PSA values as well as to diagnosis of secondary bone metastasis, which suggest a case of more severe prostate cancer. Supplementary Figure 5 shows the average attention weight across the patient population for an additional set of laboratory tests, further confirming the alignment with medical knowledge.

**Figure 4.**
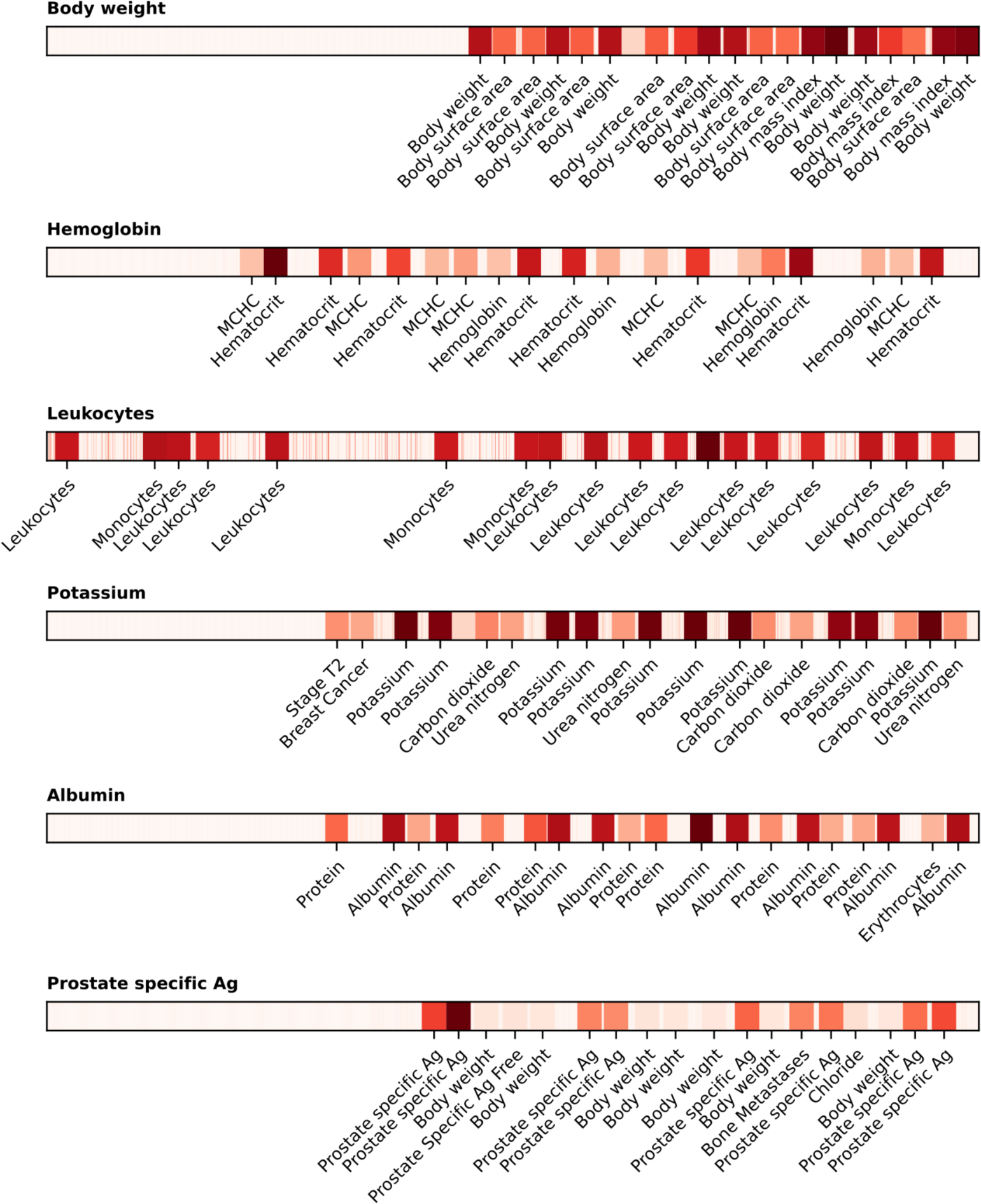
Interpretation of LIFE predictions using multi-head attention weights. Every bar corresponds to a patient timeline, with the specific laboratory test labeled at the top. The observations within the timeline are color-coded to indicate the intensity of their respective attention weights. The darker the color, the greater and more relevant the weight. The top 20 observations to which the model allocated the highest attention are identified along the x-axis. It is important to note that less relevant and unlabeled observations have been spatially compressed to conserve space. Thin red lines on some of the plots represent observations that, while not among the top 20 in terms of attention weight, still garnered sufficient attention to be highlighted on the heat map. MCHC = mean corpuscular hemoglobin; Ag = antigen.

## Discussion

This paper introduces LIFE, a novel deep learning framework for laboratory data imputation in patient clinical history of real-world EHRs. LIFE leverages time decay and multi-head attention to consume all available data in patients’ clinical histories and impute missing laboratory test values at scale. Evaluation results show that LIFE was able to impute values for all the 344 laboratory tests included in the experiments with high accuracy and outperformed the state of the art. The improvement of LIFE with respect to rule-based models (19%) is particularly significant, given that these baselines are the most prevalent model type capable of scaling to impute a large amount of laboratory data and are often utilized in RWD analysis.

This work addresses an important gap in the literature, as most existing imputation models for laboratory data were not designed for RWD, which is unique in terms of its large scale, heterogeneity, and noise. Overcoming these challenges, LIFE provides a mechanism to predict missing laboratory test values by simply requiring a laboratory test, a unit of measure, and a point in time.

Because LIFE enables laboratory data imputation at scale, it opens new possibilities for downstream applications that rely on EHRs. For example, by imputing missing values at multiple points in time throughout a patients’ history, LIFE could facilitate and improve tools for cohort selection, clinical trial matching, outcome measurement, risk stratification, clinical decision support, and causal inference.^5^ One of the most promising applications is clinical trial matching, where laboratory values are used in inclusion and exclusion criteria. Patients are often not matched to a trial because of missing data. LIFE promises to reduce this problem by increasing the amount of longitudinal data available for matching algorithms to identify patients.

To demonstrate the benefits in downstream tasks, we showed how using imputations from LIFE improved performance on detecting adverse events. This is a common application in personalized healthcare and it is crucial for enhancing clinical practice. By analyzing data from EHRs, healthcare providers can anticipate potential adverse events early. This enables timely interventions, personalized treatments, and better resource allocation, ultimately leading to improved patient outcomes.^27–29^ The inclusion of additional laboratory values enhances prediction accuracy and empowers clinicians to make informed decisions, resulting in a safer and more efficient healthcare environment.

The main component of LIFE is multi-head attention. Transformers and attention mechanisms have already been used with EHRs, commonly to train foundation models from structured EHRs and clinical notes to improve predictive modeling tasks.^22,30,31^ These models were all trained to predict masked discrete tokens and did not include laboratory values. To the best of our knowledge, LIFE is the first model for EHRs based on transformers, which (1) jointly consumes both discrete concepts and continuous values from laboratory data and (2) predicts continuous values.

Due to the attention architecture, LIFE imputations are readily interpretable. Experiments showed that the model primarily leveraged three categories of clinical features: (1) past and future values of the specified laboratory test; (2) closely related laboratory tests; and (3) other related clinical observations. Not only do these features align medically, but they also showcase the wide range of information that LIFE can employ to make predictions, confirming the initial hypothesis that incorporating supplementary EHR concepts could enhance contextualization and refinement of the imputed values.

While there is a large literature of existing techniques for dealing with missing laboratory values, they all have significant limitations when applied to RWD. For example, there are thousands of different laboratory tests that may need to be imputed in a large multi-institute dataset of EHRs, but existing machine learning methods have only been used to impute 10 to 50 laboratory tests at once. Moreover, existing methods do not utilize all of a patient’s EHR. Our results demonstrate that using the entire EHRs translates to better overall performance on both imputation and downstream detection, while also affording scalability that previous methods lack. Existing imputation models also miss an opportunity to tackle different but closely related problems in EHRs, such as standardization and outliers. Healthcare data firms frequently combine data from multiple institutions and EHR systems. Standardizing laboratory codes, units of measure, and values across these different data sources is a herculean and error-prone process that is often passed on to the data consumer. Outlier values are particularly difficult to address at scale because it is difficult to distinguish between true physiologic outliers that are secondary to disease and outliers from false data entry. For example, a white blood cell count of 300 thousand/microliter could be a result of false data entry, but it could also be a real value from a patient with leukemia. While a substantial body of research has sought to address these problems separately, solutions largely require considerable time and domain expertise. LIFE potentially improves both issues. By specifying proper queries and units of measure, we can ensure that all imputed laboratory tests follow the same standards, and we can convert existing data not following standards as well. By imputing values when there is an outlier, we can verify if that outlier is physiological (in that case it would likely be replicated by LIFE) or a false data point (LIFE would predict a completely different value).

This work comes with some limitations. Some laboratory data may be missing not at random, which means that the probability that a value is missing is related to the value itself.^32^ As an example, sometimes laboratory tests are ordered in panels, and all the tests included in the panel are performed. Consequently, when we want to impute values for a test in a panel, chances are that the whole panel would be missing. However, when validating, we mask individual laboratory values rather than entire panels. This gives the imputation models more information than they might otherwise have in real-world scenarios. This limitation is a widespread concern when evaluating laboratory data imputation and, notably, it has yet to be addressed in the existing literature.^7–9^ However, the performance of LIFE, evaluated both absolutely and in comparison to baselines, in laboratory tests such as HbA1C and PSA, typically not included in panel orders, indicates its effectiveness even for standalone tests.

A challenge for all methods for EHR imputation is that there are no standard benchmark real-world datasets to validate against. The MIMIC datasets are not suitable for our use case because they are too homogenous and do not exhibit the pathologies common to most RWD. LIFE was designed to handle and organize larger datasets aggregated from multiple institutions. For this reason, we validated the model using the Tempus data warehouse. We recognize that the subset of the database selected here, while serving as a notable illustration of RWD, consists solely of oncology patients. However, given the diverse spectrum of oncology patients included, encompassing cured, in remission, terminal, and early-stage cases, alongside the dataset’s size and the comprehensive nature of available EHRs, including all comorbidities, we believe that the described performance is sufficiently robust and generalizable. It is noteworthy that LIFE performs well across cancer indications and stages (Supplementary Table 2), adding a promising indication that it would generalize to non-oncology patient populations.

It is also crucial to underscore that imputation, while a valuable tool for handling missing data, is inherently a limited technique and should not be viewed as a substitute for actual, measured laboratory values. Utilizing imputed values in clinical practice would necessitate rigorous validation for each specific laboratory test within the context of the clinical setting.

Future works will attempt to address the aforementioned limitations, including validating LIFE on a general EHR dataset. Another direction is to provide error estimates along with the predicted laboratory values, for example using ideas from conformal prediction^33^. This would allow users to assess how confident the model is in its predictions, in addition to interpretability analysis, and how much uncertainty there is in the data. Another possibility is to extend LIFE to handle discrete concepts in addition to continuous values. This would enable the user to impute discrete laboratory results (e.g., EGFR positive or negative) and discrete observations as well (e.g., presence of a disease, medications, biomarkers). Rather than multiple systems working with different patient representations derived for different tasks, joint modeling would be capable of handling all features from EHRs at the same time. Building upon LIFE, such future work could contribute to the next generation of clinical systems that use a single-foundation model from EHRs to include millions of patients and features to effectively support the healthcare industry and clinicians.

## Methods

This section presents an overview of the LIFE architecture and a description of the evaluation design (see Figure 1).

### Dataset

We used de-identified EHRs from the Tempus data warehouse for both model training and testing. This database contains de-identified, structured EHRs of oncology patients gathered from over 350 direct data connections across approximately 2,000 healthcare institutions in the United States that order Tempus products and services. We included in the study only patients with at least one continuous laboratory value in their clinical history. This led to a dataset of 1,109,530 patients, spanning the years 2000 to 2023, comprising 658,780 females, 449,212 males, and 1,538 not declared, with a mean age of 69.1 years (standard deviation = 15.8) as of 2023.

For each patient, we aggregated general demographic details (i.e., age, sex, and race) and clinical descriptors. We included ICD-10 diagnosis codes, medications normalized to RxNorm, Current Procedural Terminology version 4 procedure codes, and vital signs and laboratory data normalized to LOINC. The vocabulary was composed of 29,454 medical concepts, leading to a dataset with an average of 727.4 records per patient.

The dataset contained 707 million laboratory values, and 4,788 distinct laboratory tests. In order to obtain meaningful predictions and not bias training because of data scarcity, we considered only the tests with at least 10,000 occurrences. This led to 344 distinct laboratory tests that were used as imputation targets in the evaluation.

The data was split into an 85% train set, 5% tuning set and 10% test set. The test set was used to generate all performance metrics reported in “Results”, whereas the tuning set was used for early stopping and hyperparameter tuning.

In addition to EHRs, the Tempus data warehouse also includes a number of variables such as cancer stage, biomarker status, and adverse events, which were manually curated by a team of clinical abstractors. Specifically, the dataset included about 273,302 patients with curated variables. While these variables were not utilized during the training of LIFE and baselines, nor in the assessment of the imputed values, they served as gold standard labels for the downstream task evaluation experiment and subtyping analyses.

### Data Processing

LIFE takes as data inputs patient EHRs and a query. EHRs include all the data used as features by the model (Figure 2A). The query specifies the details of the laboratory test to predict, including unit of measure and point in time.

Every patient record in the data warehouse is aggregated as a longitudinal sequence of medical observations from EHRs, which represent different measurements or events recorded in the patient’s medical history. Each observation consists of four fields: a code, a value, a unit of measure, and a date. The observation code is a high-level description of the observation, such as the ICD-10 C34 “Lung Cancer” code or the LOINC 2857-1 “PSA” code. The observation value is the result associated with an observation. This can be a categorical value (e.g., a positive or negative result for EGFR status) or a numerical value (e.g., a glucose level). The observation unit of measure is the unit of the continuous value. Lastly, the observation date is the point in time when the observation was recorded. Some of these fields might be unavailable depending on the observation type. For example, an ICD-10 diagnosis code observation is only composed of code and date fields, without having any associated value.

We first embed codes, units of measure, and categorical values into an embedding space which is learned during training. Observation dates are converted to the number of days from the first observation in the patient’s history. Continuous values are mapped to a uniform distribution on [0, 1] using the quantile transform,^34^ and then zero-centered by subtracting 0.5. For instance, a height value of 6’2” was first mapped to 0.97 as this height corresponds to the 97th percentile in the population, and then to 0.47 in a zero-center scale. This is done to standardize all values on the same scale, minimize the impact of outliers, and facilitate the comparison of model performance across laboratory results.

The embeddings and the continuous value are then concatenated to form a single vector for each observation. We lastly combine all the observations together into a N x E matrix, where N is the number of observations and E is the embedding size of each observation.

The query has the same structure as an observation from the EHRs: a code, a value, a unit of measure, and a date. In this case, since the value is the prediction target, it is always missing or masked. The same preprocessing of EHR observations is applied to also map the query into the same embedding space and to contextualize it in the longitudinal patient histories.

### LIFE Architecture

LIFE is a method based on self-supervised learning to process EHRs and impute missing laboratory values into a patient’s clinical history. The architecture is composed of three main blocks: a time decay module, a multi-head attention layer, and the Hadamard module (Figure 2 B-D).

The time decay module assigns different weights to each observation based on how close its date is to the query date. This architecture has been shown to outperform RNNs or CNNs when used to capture temporal information in structured EHRs.^35^ For each observation, a weight is determined as:

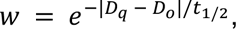

where *w* is the resulting weight, *D_q_* is the date in the query expressed in days, *D_o_* is the date of the observation in days, and *t_1/2_* is the half-life coefficient, which is the time required to decrease the relevance of the observation to one-half. We used different values for the half-life parameter to model different temporal windows. To this aim, we fed the inputs into a number of identical models but with different half-lives and concatenated their results together in the final module.

The multi-head attention layer computes similarity scores between each observation and the query, and uses them to generate patient embeddings that capture the most important features for prediction.^21^ For example, if the query is “Body Weight”, the model might pay more attention to observations related to body mass index than heart rate. Considering the context of the standard transformer architecture, keys correspond to the embedded EHR observations, while values are derived by multiplying each EHR observation with its respective time decay weight. This approach enables the architecture to amalgamate temporal and medical relevance within a single attention mechanism. The output of this layer is a batch-normalized one-dimensional embedding vector that represents the patient’s state with respect to the query.

The Hadamard module combines the output of the attention module with the query and produces a single scalar value as the prediction. This model performs a component-wise multiplication between the patient representation vector and the query vector to create a new embedding that combines the features of both inputs. It then concatenates the output of the different time decay modules and feeds the resulting vector into layers of dense neural networks. The last layer outputs a scalar value between 0 and 1, which is the quantile prediction of the query.

LIFE is trained in a self-supervised fashion by predicting a masked deleted laboratory value in a patient’s EHR (Figure 2A). This mechanism is inspired by the Bidirectional Encoder Representations from Transformers architecture,^36^ in which a transformer model is trained to interpret text by predicting a deleted word. The main difference is that LIFE does not predict a missing laboratory code but only its value since only the value is masked. Prediction is then compared to the original laboratory value via mean squared error (MSE) loss and the weights are updated via backpropagation until convergence.

### Implementation Details

All model hyperparameters were empirically tuned using the tuning set to minimize the architecture loss, while balancing training efficiency and computation time. We tested a number of configurations (e.g., data embedding space of size {128, 256, 512, 1024}; learning rate equal to {0.1, 0.01, 0.001, 0.0001}; attention embedding space of size {256, 512, 1024, 2048}; number of decay modules *D* = {1, 3, 5} and half-lives {7, 30, 60, 180, 365}). For brevity, we report only the final setting used in the results described in the evaluation. All modules were implemented in Python 3.9.16, using scikit-learn 1.2.1, pytorch 1.12.0, and pytorch-lightning 1.9.0 as machine learning libraries.

We used a 256-dimensional space to embed codes, units of measure, and categorical results. The multi-head attention layer was composed of 1024-dimensional embeddings and 8 heads. Other hyperparameters were the default set by pytorch. We used three time-decay modules with half-lives of 7, 30, and 365 days. These were chosen to simulate different clinical scenarios of inpatient visits, frequent outpatient visits, and regular outpatient visits, respectively. The Hadamard module was composed of two multilayer perceptron (MLP) networks. The first layer had 3,072 hidden units and an ReLU activation function; the second and final MLP had 1,024 hidden units and a Sigmoid activation function to output a continuous value between 0 and 1.

The model was trained using Adam optimizer, with a learning rate of 0.001 and a batch size of 128 per GPU with Distributed Data Parallelism^37^ (DDP) on 8 GPUs (effective batch size of 1024). To accommodate for the large batch size, we used accumulated gradients, which update the model parameters after accumulating gradients over several mini batches.^38^ All models were trained for 30 epochs with early stopping if the MSE of the tuning set did not improve on two successive epochs.

### Baselines

We compared LIFE with two rule-based methods (Interpolation and Nearest Lab) and three statistical or machine learning methods (MICE, MD-MTS, and BRITS). Unless otherwise specified, we implemented each method as reported in the literature or using the default configuration provided in the corresponding scikit-learn and pytorch packages.

Interpolation imputes the average value of the same laboratory test before and after the prediction date. If one of these values was not available, we simply imputed the other value. If both were missing, we imputed the distribution average for that laboratory test in the training set. Nearest Lab imputes the laboratory value which is close in time to the prediction date.

Similarly as above, if no values were available in the patient history, we imputed the distribution average in the training set. MICE is a statistical method that runs multiple regression models with each missing value being modeled conditional upon other values in the data.^6^ MICE only uses cross-sectional features at query date for imputation. If a laboratory value was not present on the query date, it was imputed using Nearest Lab. Since there is no exact implementation of MICE in Python, we used the closely related IterativeImputer method from scikit-learn.

MD-MTS is a LightGBM-based model that performed best on a recent laboratory value imputation competition using the MIMIC III dataset.^7,12^ For each laboratory test, MD-MTS builds a separate LightGBM model using a number of hand-crafted features, specifically: (1) cross-sectional laboratory data available on the same visit; (2) same laboratory values from the past and future three visits around the query date; (3) temporal information such as the number of days since the start of the patient’s history in the EHRs; and (4) summary statistics such as the minimum, mean, and maximum of the laboratory test of interest. LightGBM is a gradient boosting framework that uses tree-based learning algorithms for classification, ranking and other general machine learning tasks.

BRITS is a deep learning architecture, which directly learns missing values in a bi-directional recurrent dynamical system.^9^ The imputed values are treated as variables of a RNN graph and, similarly to LIFE, are updated during the backpropagation loop. We trained BRITS in parallel using DDP on 8 GPUs with a batch size of 32 per GPU and Adam optimizer with the default learning rate. Acknowledging that LIFE benefited from larger batch sizes owing to the large amount of noise in real-world EHRs, we attempted to do this with BRITS as well, but this did not improve results.

### Experimental Design

We conducted a number of experiments to assess LIFE’s effectiveness in imputing clinically meaningful laboratory data.

First, we trained the model on all 344 laboratory tests included in the dataset. For each patient in the test set (n=110,953), we selected from their clinical history one random laboratory value from these 344 options and deleted it. We then used LIFE to predict the missing values. We finally grouped patients with the same deleted laboratory test together and measured average MAE per laboratory test.

We then compared LIFE’s performance to all baselines on a subset of laboratory tests. Due to the baselines’ less scalable design, we focused on 25 common laboratory tests, which is approximately the number of tests baselines were evaluated in the literature. For each laboratory test, we randomly selected 10,000 patients who had at least one observation of that test in their EHRs. For each test patient, we deleted one observation of the laboratory test of interest and used it as the prediction target. We then compared each models’ ability to predict this value given the rest of the patient’s EHRs and the query for the laboratory test. We chose a sample size of 10,000 patients per laboratory test based on a power calculation to ensure the statistical significance of our results using two criteria: (1) statistically significant was defined as p < 0.01 calculated via a two-sample t-test and (2) MAE effect size would be 0.01. For some laboratory tests, there were not enough patients available in the test set. In this case, we used the maximum available number of patients.

We also conducted a series of supplementary analyses to assess the performance of LIFE when applied to specific clinical scenarios. Specifically: (1) we evaluated how effectively the model performs at different points in the patient’s timeline, shedding light on its potential to predict future laboratory values; (2) we analyzed how the magnitude of laboratory values impacted LIFE’s performance; (3) we evaluated whether LIFE could categorize predicted values as “abnormal” by measuring the distance between predicted continuous values and laboratory test median and (4) we delved into the robustness of the model across various levels of cancer severity, to verify performances in early-stage and benign diseases, which would indicate that LIFE may generalize well when applied to non-oncology patients. These analyses are described further in the appendix.

We then measured the performances of imputed laboratory values on the downstream task of detecting adverse events from laboratory data. To do this, we selected nine adverse events that are frequent in oncology patients using curated variables available in the Tempus data warehouse. For each adverse event, we identified a cohort of patients from the test set with that adverse event and the same number of random patients without it. Afterward, we performed sub-sampling on all cohorts to match the size of the smallest cohort, resulting in each cohort consisting of 6,932 patients. We then used LIFE and all baselines to impute values for the same 25 laboratory tests at the time of the corresponding adverse event. For each adverse event, we then used a 80/20 split to train and evaluate a logistic regression classifier which takes as input the feature vector composed of the 25 imputed laboratory values and output if the patients had that adverse event or not. We evaluated averaged performances of all models and across all patients in terms of AUC-PR.

Lastly, we generated plots to visualize attention weights produced by LIFE’s multi-head attention layer, in order to interpret the imputation predictions. We randomly selected six patients with different laboratory tests evaluated during our first experiment. Each patient’s timeline was represented as a band with the laboratory of interest labeled at the top. The observations within these timelines were color-coded based on the intensity of their attention weights (average across all heads), allowing for an easy assessment of the model’s focus. To further enhance the interpretability of the plot, we specifically labeled only the 20 observations with the highest attention weights along the x-axis. The less relevant observations were collapsed in the interest of conserving space and improving the overall readability of the plot. To illustrate that LIFE’s interpretability is scalable we also conducted a supplementary analysis in which we calculated the average attention paid to different features across the entire patient population for an additional subset of tests.

## Author Contributions

S.P.H. and R.M. conceived and designed the work. S.P.H. conducted the research and the experimental evaluation, provided clinical validation of the results and drafted the manuscript. R.M. created the dataset, supervised and supported the research, and substantially edited the manuscript. C.C. advised on methodological choices, contributed to the experimental evaluation and critically revised the manuscript. D.M.V. critically revised the manuscript. E.T.M. and M.C.S. supported the research and revised the manuscript. All the authors gave final approval of the completed manuscript version and are accountable for all aspects of the work.

## Data Availability

Deidentified data used in the research was collected in a real-world health care setting and is subject to controlled access for privacy and proprietary reasons. When possible, derived data supporting the findings of this study have been made available within the paper and its figures and tables.

## Code Availability

Requests for source code are subject to review by Tempus and can be directed to the lead author.

## Supplementary Material

**Supplementary Figure 1.**
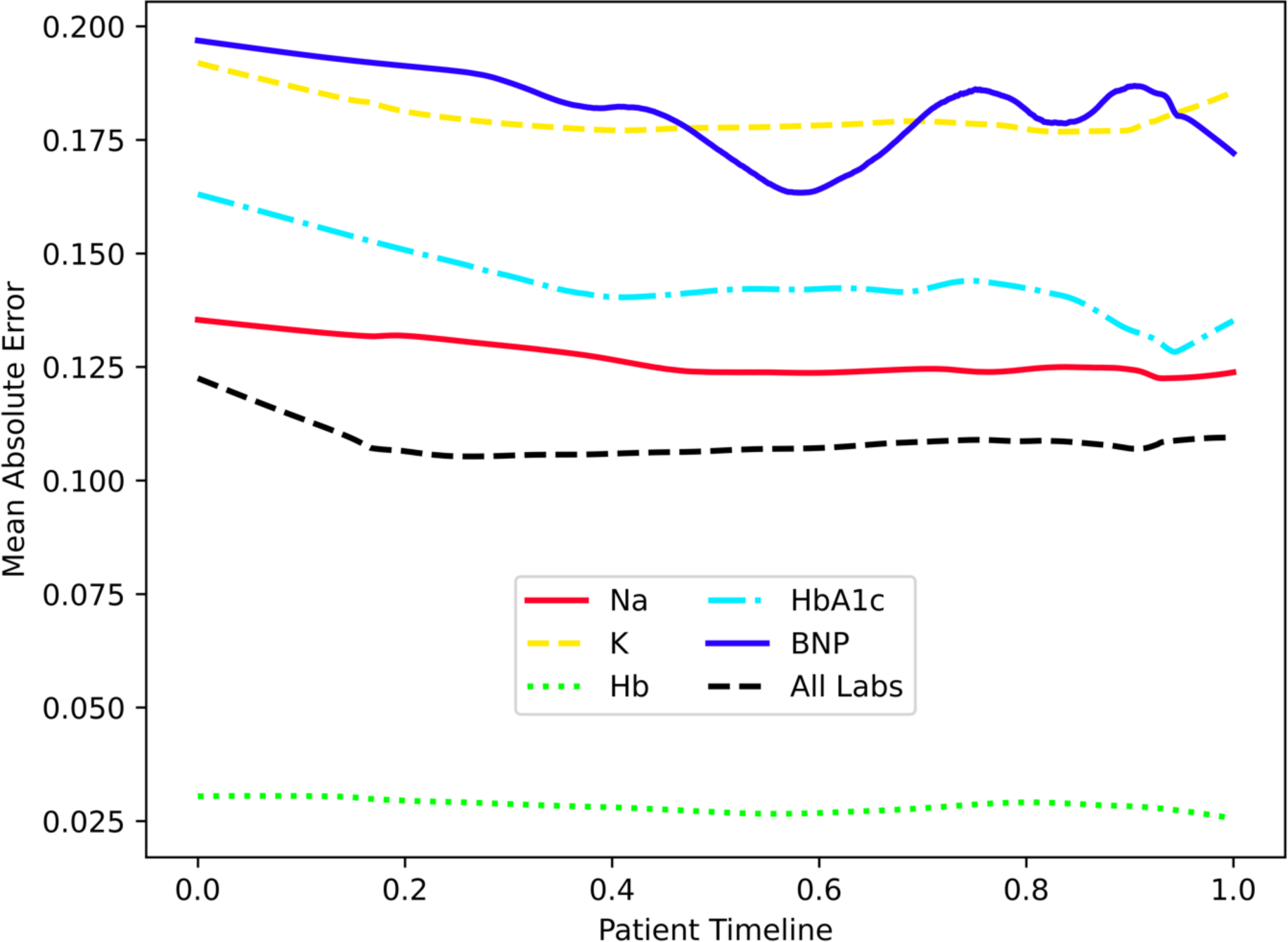
LIFE’s performance with respect to prediction timing within a patient’s history. Laboratory values imputed by LIFE remain unaffected by the timing of predictions within a patient’s history. Specifically, LIFE exhibits strong performance when imputing laboratory values at the beginning, middle, or end of the patient’s known history. The x-axis represents the normalized patient timeline, with 0 denoting the first record and 1 indicating the last. The y-axis represents the mean absolute error of the laboratory values predicted at various points along the timeline. Plot lines were generated using Locally Estimated Scatterplot Smoothing (LOESS), utilizing performance data extracted from the test set.

**Supplementary Figure 2.**
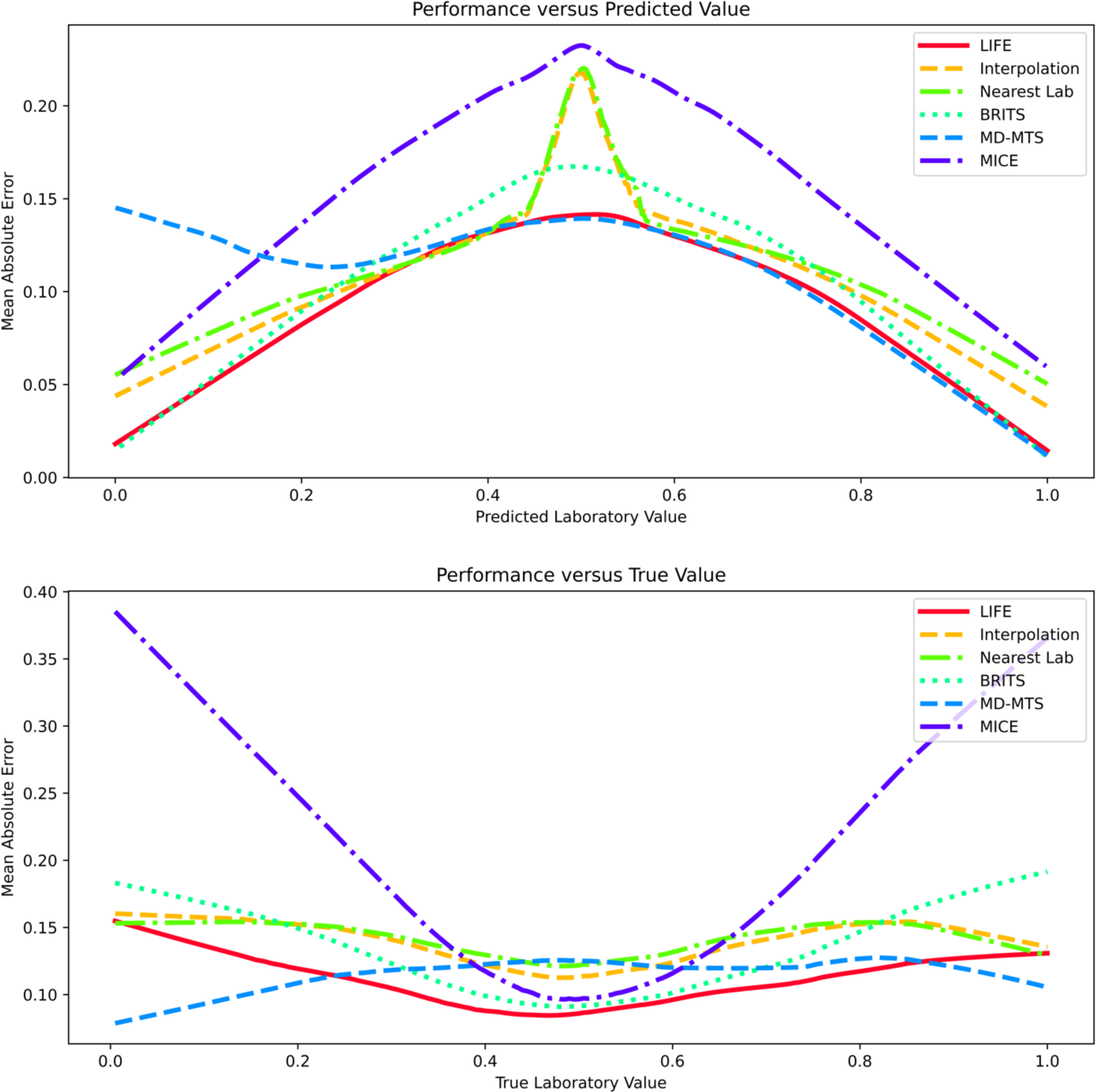
Model error as a function of laboratory value magnitude for LIFE and baselines. Imputation model performance varies depending on the extremity of the predicted laboratory value. In terms of *predicted* laboratory values (see the upper panel), LIFE exhibited enhanced accuracy for extreme values and diminished accuracy for median values. We hypothesize that, in order to minimize the mean squared error (MSE), models predict a value near the median when there is a high level of uncertainty in the prediction. As a result, predicted values near the median have a higher error. The rule-based baselines exhibited particularly poor performance for predicted values of 0.5. This can be attributed to the absence of previous or future laboratory values for these models to utilize, leading them to predict a default value of 0.5. We observed the exact opposite for *true* laboratory values (see the lower panel), where LIFE had diminished accuracy for extreme values and enhanced accuracy for central or median values. *True* laboratory values are usually extreme as a result of acute illness. For example, a patient’s white blood cell count will spike if they develop an infection. Because acute illness happens partly at random – due to, for example, exposure to a deadly pathogen – all models have a harder time predicting them based on the patient’s baseline past and future values. The trends for both true and predicted values resembled those observed in the baseline models, but we noted that LIFE tended to mitigate these behaviors. The graphical lines were generated using Locally Estimated Scatterplot Smoothing (LOESS), which employed performance data derived from the test set.

**Supplementary Figure 3.**
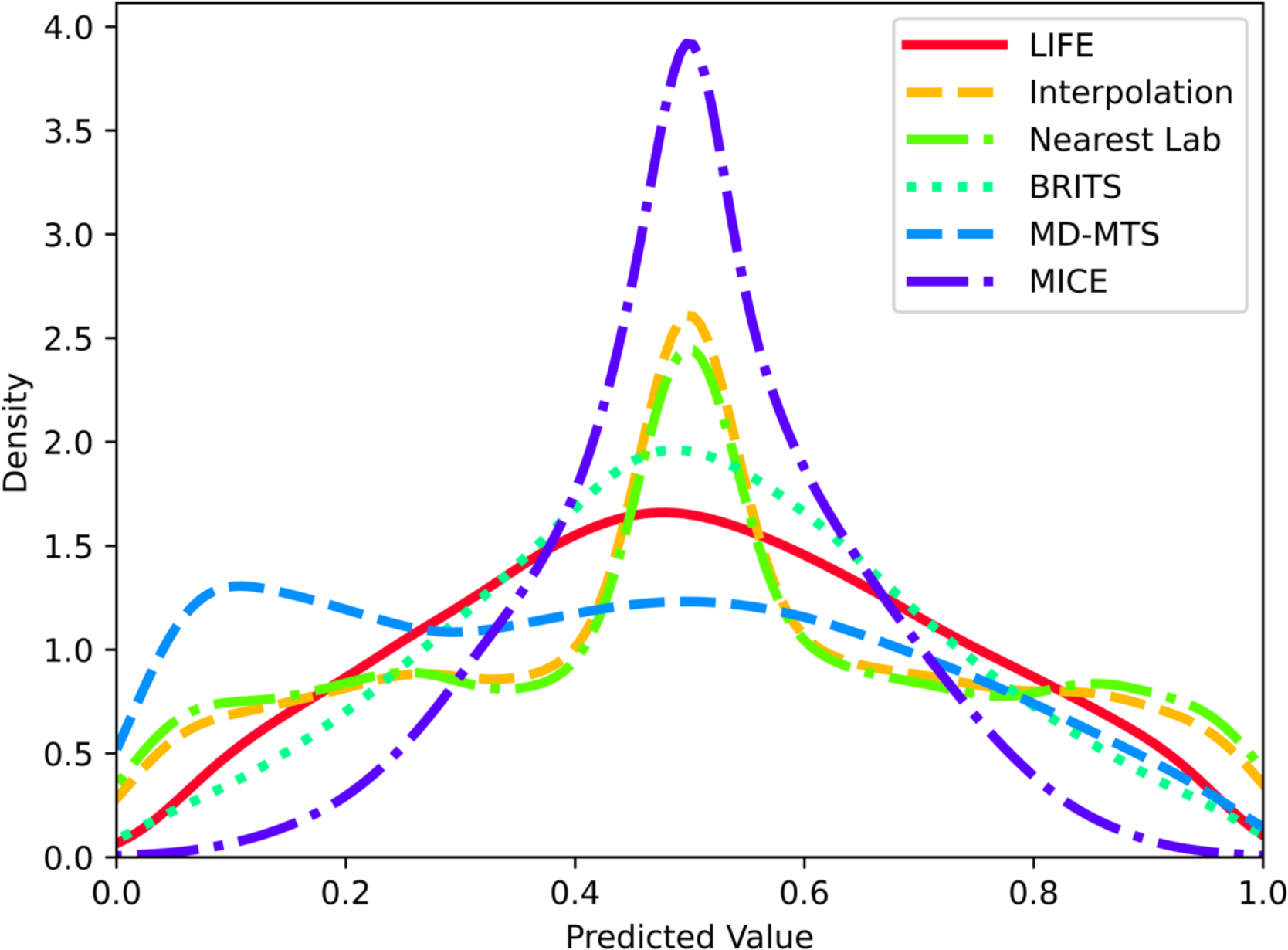
Distribution of predicted laboratory values. All models exhibit a bias toward predicting central or median values. When the model does not have enough data to make an accurate prediction, we hypothesize that it predicts a value near the median to minimize the mean squared error. LIFE’s centrality bias is similar to other neural network baselines like BRITS. As expected, the Nearest Lab and Interpolation show a fairly uniform distribution with a peak around 0.5, corresponding to the fact that these models predict 0.5 when there are no past or future laboratory values to use. The graphical lines were generated using Locally Estimated Scatterplot Smoothing (LOESS), employing prediction data derived from the test set.

**Supplementary Figure 4.**
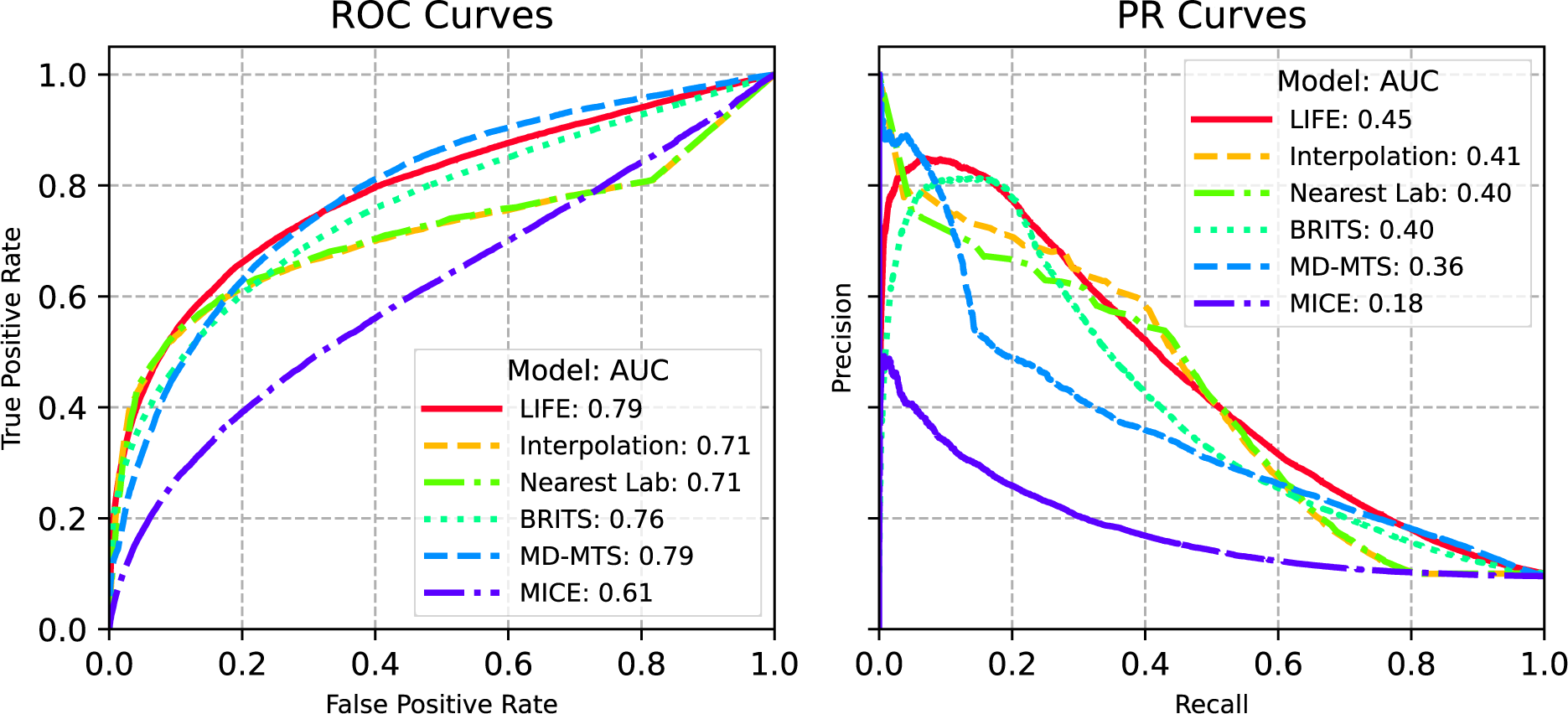
LIFE and baseline performances when imputing abnormal laboratory values. Laboratory values imputed by LIFE exhibited the best performance when categorized as “normal” or “abnormal”. Abnormal values were defined as those above the 95th percentile or below the 5th percentile for the population of interest. We classified predicted laboratory values as abnormal when the absolute difference between the predicted continuous value and the 50th percentile exceeded a set threshold. For each model, the threshold was varied to produce receiver operating characteristic and precision recall plots. A simpler approach to predicting abnormal values would be to forgo the threshold and simply predict an abnormal true value if the predicted continuous value was above the 95th percentile or below the 5th percentile. This corresponds to a single point on the above PR and ROC curves, where the fixed threshold is 0.45. We opted to vary the threshold for categorizing predicted laboratory values as abnormal for two main reasons. First, using a fixed threshold makes it challenging to compare the performance of different models. This is because each model exhibits unique sensitivity and specificity characteristics when evaluated at a single, constant threshold. By allowing the threshold to vary, we generate PR and ROC plots that offer a more robust basis for cross-model comparison. Second, the likelihood that a predicted laboratory value is genuinely abnormal does not strictly depend on whether the predicted value falls above the 95th percentile or below the 5th percentile. In practice, the probability of a result being abnormal increases gradually as the predicted value deviates more significantly from the median. Varying the threshold allows us to better capture and represent this continuous relationship.

**Supplementary Figure 5.**
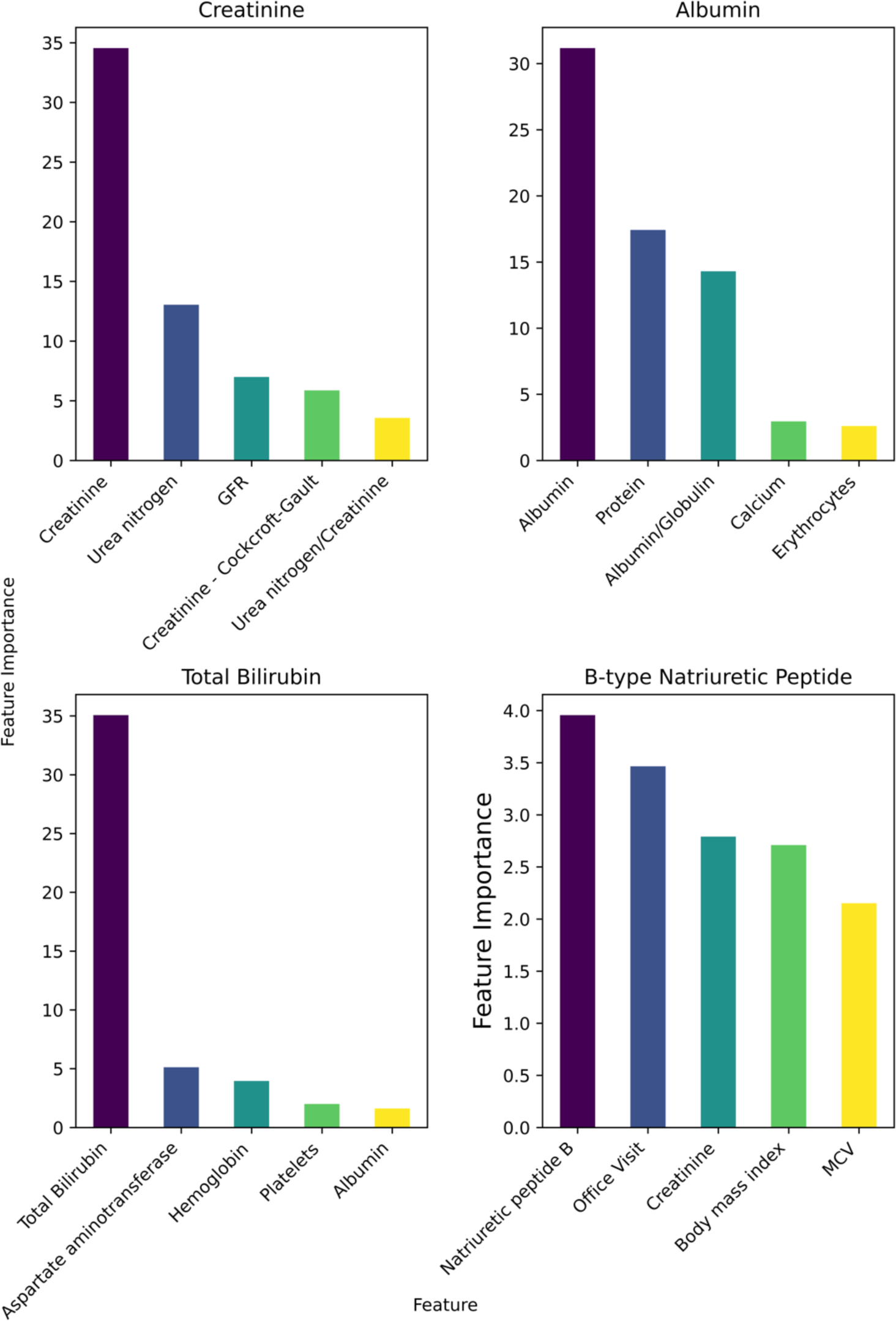
Top five features in LIFE’s imputations for a selection of laboratory tests. Feature importance was determined by computing the average attention allocated to each type of observation (e.g., total bilirubin, body mass index) across all test set patients for whom the corresponding laboratory values were predicted. In all cases, the predominant feature influencing these predictions was the laboratory result at other time points, followed by other relevant laboratory results. For instance, when forecasting albumin levels, the patient’s protein level emerged as the second most significant feature, aligning with the understanding that albumin is the primary protein present in blood. Interestingly, B-type Natriuretic Peptide (BNP) relied more heavily on observations beyond its own past and future values. This is likely due to the infrequent ordering of this laboratory test, compelling the model to depend on other observations for making predictions.

**Supplementary Table 1.** LIFE results on 344 laboratory tests in terms of mean absolute error (MAE). For each patient in the test set, the laboratory to predict was chosen at random from the pool of recorded values. The selected laboratory test was masked and used to validate the model’s prediction. The average MAE is equal to 0.15. In the first column of the table the laboratory tests LOINC code is specified in parentheses.

| Laboratory Test | MAE |
| --- | --- |
| 25-Hydroxycalciferol [Mass/volume] in Serum or Plasma (49054-0) | 0.03 |
| 25-Hydroxyvitamin D3+25-Hydroxyvitamin D2 [Mass/volume] in Serum or Plasma (62292-8) | 0.21 |
| Abnormal lymphocytes/100 leukocytes in Blood (30413-9) | 0.39 |
| Activated clotting time (ACT) of Blood by Coagulation assay (3184-9) | 0.16 |
| Alanine aminotransferase [Enzymatic activity/volume] in Serum or Plasma (1742-6) | 0.13 |
| Alanine aminotransferase [Enzymatic activity/volume] in Serum or Plasma by No addition of P-5'-P (1744-2) | 0.19 |
| Alanine aminotransferase [Enzymatic activity/volume] in Serum or Plasma by With P-5'-P (1743-4) | 0.10 |
| Albumin [Mass/volume] in Serum or Plasma (1751-7) | 0.10 |
| Albumin [Mass/volume] in Serum or Plasma by Bromocresol green (BCG) dye binding method (61151-7) | 0.13 |
| Albumin [Mass/volume] in Serum or Plasma by Bromocresol purple (BCP) dye binding method (61152-5) | 0.11 |
| Albumin [Mass/volume] in Serum or Plasma by Electrophoresis (2862-1) | 0.12 |
| Albumin/Globulin [Mass Ratio] in Serum or Plasma (1759-0) | 0.13 |
| Albumin/Protein.total in Serum or Plasma (35706-1) | 0.09 |
| Albumin/Protein.total in Serum or Plasma by Electrophoresis (13980-8) | 0.14 |
| Albumin/Protein.total in Urine by Electrophoresis (13992-3) | 0.33 |
| Alkaline phosphatase [Enzymatic activity/volume] in Serum or Plasma (6768-6) | 0.12 |
| Alpha 1 globulin [Mass/volume] in Serum or Plasma by Electrophoresis (2865-4) | 0.12 |
| Alpha 1 globulin [Mass/volume] in Urine by Electrophoresis (9734-5) | 0.23 |
| Alpha 1 globulin/Protein.total in Serum or Plasma by Electrophoresis (13978-2) | 0.18 |
| Alpha 1 globulin/Protein.total in Urine by Electrophoresis (13990-7) | 0.29 |
| Alpha 2 globulin [Mass/volume] in Serum or Plasma by Electrophoresis (2868-8) | 0.20 |
| Alpha 2 globulin [Mass/volume] in Urine by Electrophoresis (38190-5) | 0.06 |
| Alpha 2 globulin/Protein.total in 24 hour Urine by Electrophoresis (13987-3) | 0.60 |
| Alpha 2 globulin/Protein.total in Serum or Plasma by Electrophoresis (13981-6) | 0.24 |
| Alpha-1-Fetoprotein [Mass/volume] in Serum or Plasma (1834-1) | 0.10 |
| Alpha-1-fetoprotein.tumor marker [Mass/volume] in Serum or Plasma (53962-7) | 0.18 |
| Ammonia [Mass/volume] in Plasma (22763-7) | 0.36 |
| Amylase [Enzymatic activity/volume] in Serum or Plasma (1798-8) | 0.29 |
| Amylase [Enzymatic activity/volume] in Serum, Plasma or Blood (77146-9) | 0.19 |
| Anion gap 3 in Serum or Plasma (10466-1) | 0.18 |
| Anion gap 4 in Serum or Plasma (1863-0) | 0.15 |
| Anion gap in Blood (41276-7) | 0.11 |
| Anion gap in Serum or Plasma (33037-3) | 0.13 |
| Aspartate aminotransferase [Enzymatic activity/volume] in Serum or Plasma (1920-8) | 0.12 |
| Aspartate aminotransferase [Enzymatic activity/volume] in Serum or Plasma by With P-5'-P (30239-8) | 0.28 |
| Band form neutrophils [# /volume] in Blood (26507-4) | 0.10 |
| Band form neutrophils [# /volume] in Blood by Automated count (30229-9) | 0.06 |
| Band form neutrophils/100 leukocytes in Blood (26508-2) | 0.15 |
| Band form neutrophils/100 leukocytes in Blood by Manual count (764-1) | 0.18 |
| Base excess in Arterial blood by calculation (1925-7) | 0.17 |
| Basophils [# /volume] in Blood (26444-0) | 0.13 |
| Basophils [# /volume] in Blood by Automated count (704-7) | 0.14 |
| Basophils [# /volume] in Blood by Manual count (705-4) | 0.15 |
| Basophils+Eosinophils+Monocytes [# /volume] in Blood (32349-3) | 0.13 |
| Basophils+Eosinophils+Monocytes/100 leukocytes in Blood (32350-1) | 0.10 |
| Basophils/100 leukocytes in Blood (30180-4) | 0.14 |
| Basophils/100 leukocytes in Blood by Automated count (706-2) | 0.15 |
| Basophils/100 leukocytes in Blood by Manual count (707-0) | 0.14 |
| Beta 1 globulin [Mass/volume] in Serum or Plasma by Electrophoresis (32730-4) | 0.06 |
| Beta 1 globulin/Protein.total in Serum or Plasma by Electrophoresis (32732-0) | 0.30 |
| Beta 2 globulin [Mass/volume] in Serum or Plasma by Electrophoresis (32731-2) | 0.17 |
| Beta globulin [Mass/volume] in Serum or Plasma by Electrophoresis (2871-2) | 0.17 |
| Beta globulin/Protein.total in Serum or Plasma by Electrophoresis (13982-4) | 0.11 |
| Beta hydroxybutyrate [Moles/volume] in Serum or Plasma (6873-4) | 0.27 |
| Beta-2-Microglobulin [Mass/volume] in Serum or Plasma (1952-1) | 0.15 |
| Bicarbonate [Moles/volume] in Arterial blood (1960-4) | 0.18 |
| Bicarbonate [Moles/volume] in Plasma (1962-0) | 0.14 |
| Bicarbonate [Moles/volume] in Serum or Plasma (1963-8) | 0.20 |
| Bicarbonate [Moles/volume] in Venous blood (14627-4) | 0.25 |
| Bilirubin.conjugated [Mass/volume] in Serum or Plasma (15152-2) | 0.13 |
| Bilirubin.direct [Mass/volume] in Serum or Plasma (1968-7) | 0.16 |
| Bilirubin.indirect [Mass/volume] in Serum or Plasma (1971-1) | 0.12 |
| Bilirubin.total [Mass/volume] in Serum or Plasma (1975-2) | 0.15 |
| Bilirubin.total [Presence] in Urine (1977-8) | 0.13 |
| Blasts [# /volume] in Blood by Manual count (708-8) | 0.08 |
| Blasts/100 leukocytes in Blood (26446-5) | 0.03 |
| Blasts/100 leukocytes in Blood by Manual count (709-6) | 0.05 |
| Body height (50373000) | 0.06 |
| Body mass index (60621009) | 0.21 |
| Body surface area (301898006) | 0.10 |
| Body temperature measure (363816005) | 0.22 |
| Body weight (27113001) | 0.05 |
| C reactive protein [Mass/volume] in Serum or Plasma (1988-5) | 0.18 |
| Calcidiol [Mass/volume] in Serum or Plasma (1989-3) | 0.21 |
| Calcium [Mass/volume] corrected for albumin in Serum or Plasma (46099-8) | 0.17 |
| Calcium [Mass/volume] in Serum or Plasma (17861-6) | 0.16 |
| Calcium [Moles/volume] corrected for total protein in Blood (47597-0) | 0.01 |
| Calcium [Moles/volume] in Serum or Plasma (2000-8) | 0.12 |
| Calcium.ionized [Mass/volume] in Blood (38230-9) | 0.34 |
| Calcium.ionized [Mass/volume] in Serum or Plasma (17863-2) | 0.15 |
| Calcium.ionized [Moles/volume] adjusted to pH 7.4 in Blood (47598-8) | 0.14 |
| Calcium.ionized [Moles/volume] in Blood (1994-3) | 0.48 |
| Calcium.ionized [Moles/volume] in Blood by Ion-selective membrane electrode (ISE) (47596-2) | 0.16 |
| Calcium.ionized [Moles/volume] in Serum or Plasma (1995-0) | 0.17 |
| Cancer Ag 125 [Presence] in Serum or Plasma (2006-5) | 0.18 |
| Cancer Ag 125 [Units/volume] in Serum or Plasma (10334-1) | 0.12 |
| Cancer Ag 15-3 [Units/volume] in Serum or Plasma (6875-9) | 0.08 |
| Cancer Ag 19-9 [Units/volume] in Serum or Plasma (24108-3) | 0.14 |
| Cancer Ag 27-29 [Units/volume] in Serum or Plasma (17842-6) | 0.10 |
| Cancer related large scale gene targeted mutation analysis in Blood or Tissue by Molecular genetics method (73977-1) | 0.25 |
| Carbon dioxide [Partial pressure] in Arterial blood (2019-8) | 0.03 |
| Carbon dioxide, total [Moles/volume] in Blood (20565-8) | 0.16 |
| Carbon dioxide, total [Moles/volume] in Serum or Plasma (2028-9) | 0.15 |
| Carbon dioxide, total [Moles/volume] in Venous blood (2027-1) | 0.18 |
| Carcinoembryonic Ag [Mass/volume] in Serum or Plasma (2039-6) | 0.14 |
| Chloride [Moles/volume] in Blood (2069-3) | 0.13 |
| Chloride [Moles/volume] in Capillary blood (51590-8) | 0.19 |
| Chloride [Moles/volume] in Serum or Plasma (2075-0) | 0.13 |
| Cholesterol [Mass/volume] in Serum or Plasma (2093-3) | 0.10 |
| Cholesterol in HDL [Mass/volume] in Serum or Plasma (2085-9) | 0.09 |
| Cholesterol in LDL [Mass/volume] in Serum or Plasma by Direct assay (18262-6) | 0.11 |
| Cholesterol in LDL [Mass/volume] in Serum or Plasma by calculation (13457-7) | 0.13 |
| Cholesterol in VLDL [Mass/volume] in Serum or Plasma by calculation (13458-5) | 0.05 |
| Cholesterol non HDL [Mass/volume] in Serum or Plasma (43396-1) | 0.11 |
| Cholesterol.total/Cholesterol in HDL [Mass Ratio] in Serum or Plasma (9830-1) | 0.17 |
| Choriogonadotropin [Units/volume] in Serum or Plasma (19080-1) | 0.12 |
| Choriogonadotropin.beta subunit [Units/volume] in Serum or Plasma (21198-7) | 0.05 |
| Chromogranin A [Mass/volume] in Serum or Plasma (9811-1) | 0.13 |
| Cobalamin (Vitamin B12) [Mass/volume] in Serum or Plasma (2132-9) | 0.21 |
| Corticotropin [Mass/volume] in Plasma (2141-0) | 0.04 |
| Cortisol [Mass/volume] in Serum or Plasma (2143-6) | 0.22 |
| Creatine kinase [Enzymatic activity/volume] in Serum or Plasma (2157-6) | 0.20 |
| Creatine kinase.MB/Creatine kinase.total in Serum or Plasma (20569-0) | 0.13 |
| Creatinine [Mass/time] in 24 hour Urine (2162-6) | 0.15 |
| Creatinine [Mass/volume] in Blood (38483-4) | 0.19 |
| Creatinine [Mass/volume] in Serum or Plasma (2160-0) | 0.10 |
| Creatinine [Mass/volume] in Urine (2161-8) | 0.10 |
| Creatinine renal clearance in 24 hour Urine and Serum or Plasma (2164-2) | 0.02 |
| Creatinine renal clearance in Urine and Serum or Plasma collected for unspecified duration (33558-8) | 0.09 |
| Creatinine renal clearance predicted by Cockcroft-Gault formula (35591-7) | 0.08 |
| Diastolic blood pressure (271650006) | 0.22 |
| Eosinophils [# /volume] in Blood (26449-9) | 0.09 |
| Eosinophils [# /volume] in Blood by Automated count (711-2) | 0.11 |
| Eosinophils [# /volume] in Blood by Manual count (712-0) | 0.14 |
| Eosinophils/100 leukocytes in Blood (26450-7) | 0.10 |
| Eosinophils/100 leukocytes in Blood by Automated count (713-8) | 0.10 |
| Eosinophils/100 leukocytes in Blood by Manual count (714-6) | 0.22 |
| Epithelial cells.squamous [# /area] in Urine sediment by Automated count (33219-7) | 0.19 |
| Erythrocyte distribution width [Entitic volume] (30384-2) | 0.10 |
| Erythrocyte distribution width [Entitic volume] by Automated count (21000-5) | 0.10 |
| Erythrocyte distribution width [Ratio] (30385-9) | 0.12 |
| Erythrocyte distribution width [Ratio] by Automated count (788-0) | 0.11 |
| Erythrocyte sedimentation rate (30341-2) | 0.24 |
| Erythrocyte sedimentation rate by Westergren method (4537-7) | 0.20 |
| Erythrocytes [# /area] in Urine sediment by Automated count (46419-8) | 0.25 |
| Erythrocytes [# /volume] in Blood (26453-1) | 0.07 |
| Erythrocytes [# /volume] in Blood by Automated count (789-8) | 0.10 |
| Erythropoietin (EPO) [Units/volume] in Serum or Plasma (15061-5) | 0.18 |
| Estradiol (E2) [Mass/volume] in Serum or Plasma (2243-4) | 0.11 |
| Ethanol [Mass/volume] in Serum or Plasma (5643-2) | 0.17 |
| Fasting glucose [Mass/volume] in Serum or Plasma (1558-6) | 0.07 |
| Ferritin [Mass/volume] in Serum or Plasma (2276-4) | 0.16 |
| Fibrin D-dimer FEU [Mass/volume] in Platelet poor plasma (48065-7) | 0.13 |
| Fibrinogen [Mass/volume] in Platelet poor plasma by Coagulation assay (3255-7) | 0.17 |
| Folate [Mass/volume] in Serum or Plasma (2284-8) | 0.27 |
| Follitropin [Units/volume] in Serum or Plasma (15067-2) | 0.19 |
| Gamma globulin [Mass/volume] in Serum or Plasma by Electrophoresis (2874-6) | 0.08 |
| Gamma globulin/Protein.total by Electrophoresis in Urine collected for unspecified duration (17817-8) | 0.30 |
| Gamma globulin/Protein.total in Serum or Plasma by Electrophoresis (13983-2) | 0.25 |
| Gamma glutamyl transferase [Enzymatic activity/volume] in Serum or Plasma (2324-2) | 0.10 |
| Globulin [Mass/volume] in Serum (2336-6) | 0.07 |
| Globulin [Mass/volume] in Serum by calculation (10834-0) | 0.05 |
| Globulin [Mass/volume] in Urine by Electrophoresis (49047-4) | 0.09 |
| Globulin [Mass/volume] in Urine by calculation (18320-2) | 0.29 |
| Glomerular filtration rate/1.73 sq M.predicted [Volume Rate/Area] in Serum or Plasma by Creatinine-based formula (MDRD) (33914-3) | 0.14 |
| Glomerular filtration rate/1.73 sq M.predicted [Volume Rate/Area] in Serum, Plasma or Blood (69405-9) | 0.11 |
| Glomerular filtration rate/1.73 sq M.predicted [Volume Rate/Area] in Serum, Plasma or Blood by Creatinine-based formula (CKD-EPI) (62238-1) | 0.10 |
| Glomerular filtration rate/1.73 sq M.predicted among blacks [Volume Rate/Area] in Serum, Plasma or Blood by Creatinine-based formula (MDRD) (48643-1) | 0.07 |
| Glomerular filtration rate/1.73 sq M.predicted among females [Volume Rate/Area] in Serum, Plasma or Blood by Creatinine-based formula (MDRD) (50044-7) | 0.10 |
| Glomerular filtration rate/1.73 sq M.predicted among non-blacks [Volume Rate/Area] in Serum, Plasma or Blood by Creatinine-based formula (CKD-EPI) (88294-4) | 0.06 |
| Glomerular filtration rate/1.73 sq M.predicted among non-blacks [Volume Rate/Area] in Serum, Plasma or Blood by Creatinine-based formula (MDRD) (48642-3) | 0.06 |
| Glucose [Mass/volume] in Blood (2339-0) | 0.16 |
| Glucose [Mass/volume] in Blood by Automated test strip (2340-8) | 0.18 |
| Glucose [Mass/volume] in Capillary blood (32016-8) | 0.19 |
| Glucose [Mass/volume] in Capillary blood by Glucometer (41653-7) | 0.17 |
| Glucose [Mass/volume] in Serum or Plasma (2345-7) | 0.17 |
| Glucose [Mass/volume] in Urine (2350-7) | 0.21 |
| Glucose mean value [Mass/volume] in Blood Estimated from glycated hemoglobin (27353-2) | 0.11 |
| Granulocytes [# /volume] in Blood (30394-1) | 0.09 |
| Granulocytes [# /volume] in Blood by Automated count (20482-6) | 0.07 |
| Granulocytes/100 leukocytes in Blood (30395-8) | 0.08 |
| Granulocytes/100 leukocytes in Blood by Automated count (19023-1) | 0.05 |
| Haptoglobin [Mass/volume] in Serum or Plasma (4542-7) | 0.32 |
| Hematocrit [Volume Fraction] of Blood (20570-8) | 0.06 |
| Hematocrit [Volume Fraction] of Blood by Automated count (4544-3) | 0.07 |
| Hematocrit [Volume Fraction] of Capillary blood (42908-4) | 0.04 |
| Hemoglobin A1c/Hemoglobin.total in Blood (4548-4) | 0.12 |
| Hemoglobin A1c/Hemoglobin.total in Blood by IFCC protocol (59261-8) | 0.05 |
| Hemoglobin A1c/Hemoglobin.total in Blood by calculation (17855-8) | 0.12 |
| Hemoglobin [Mass/volume] in Blood (718-7) | 0.07 |
| Hemoglobin [Mass/volume] in Blood by calculation (20509-6) | 0.06 |
| Hemoglobin saturation with oxygen (103228002) | 0.21 |
| Hemolysis index of Serum or Plasma (46424-8) | 0.07 |
| Hepatitis C virus Ab Signal/Cutoff in Serum or Plasma by Immunoassay (48159-8) | 0.26 |
| INR in Blood by Coagulation assay (34714-6) | 0.28 |
| INR in Platelet poor plasma by Coagulation assay (6301-6) | 0.25 |
| INR in Platelet poor plasma or blood by Coagulation assay (38875-1) | 0.24 |
| IgA [Mass/volume] in Serum or Plasma (2458-8) | 0.12 |
| IgG [Mass/volume] in Serum or Plasma (2465-3) | 0.12 |
| IgM [Mass/volume] in Serum or Plasma (2472-9) | 0.11 |
| Immature granulocytes [# /volume] in Blood (51584-1) | 0.11 |
| Immature granulocytes [# /volume] in Blood by Automated count (53115-2) | 0.14 |
| Immature granulocytes [Presence] in Blood by Automated count (34165-1) | 0.14 |
| Immature granulocytes/100 leukocytes in Blood (38518-7) | 0.14 |
| Immature granulocytes/100 leukocytes in Blood by Automated count (71695-1) | 0.13 |
| Immature reticulocytes/Reticulocytes.total in Blood (33516-6) | 0.21 |
| Iron [Mass/volume] in Serum or Plasma (2498-4) | 0.12 |
| Iron binding capacity [Mass/volume] in Serum or Plasma (2500-7) | 0.12 |
| Iron binding capacity.unsaturated [Mass/volume] in Serum or Plasma (2501-5) | 0.07 |
| Iron saturation [Mass Fraction] in Serum or Plasma (2502-3) | 0.07 |
| Kappa light chains [Mass/volume] in Serum or Plasma (11050-2) | 0.10 |
| Kappa light chains.free [Mass/volume] in Serum (36916-5) | 0.11 |
| Kappa light chains.free/Lambda light chains.free [Mass Ratio] in Serum (48378-4) | 0.15 |
| Kappa light chains/Lambda light chains [Mass Ratio] in Serum (15189-4) | 0.19 |
| Lactate [Moles/volume] in Blood (32693-4) | 0.34 |
| Lactate [Moles/volume] in Serum or Plasma (2524-7) | 0.16 |
| Lactate dehydrogenase [Enzymatic activity/volume] in Serum or Plasma (2532-0) | 0.13 |
| Lactate dehydrogenase [Enzymatic activity/volume] in Serum or Plasma by Lactate to pyruvate reaction (14804-9) | 0.14 |
| Lambda light chains.free [Mass/volume] in Serum or Plasma (33944-0) | 0.11 |
| Leukocytes [# /area] in Urine sediment by Automated count (46702-7) | 0.23 |
| Leukocytes [# /volume] corrected for nucleated erythrocytes in Blood (12227-5) | 0.03 |
| Leukocytes [# /volume] corrected for nucleated erythrocytes in Blood by Automated count (33256-9) | 0.04 |
| Leukocytes [# /volume] in Blood (26464-8) | 0.10 |
| Leukocytes [# /volume] in Blood by Automated count (6690-2) | 0.08 |
| Leukocytes [# /volume] in Blood by Manual count (804-5) | 0.11 |
| Leukocytes [# /volume] in Urine (30405-5) | 0.34 |
| Lipase [Enzymatic activity/volume] in Serum or Plasma (3040-3) | 0.16 |
| Lutropin [Units/volume] in Serum or Plasma (10501-5) | 0.14 |
| Lymphocytes [# /volume] in Blood (26474-7) | 0.08 |
| Lymphocytes [# /volume] in Blood by Automated count (731-0) | 0.11 |
| Lymphocytes [# /volume] in Blood by Manual count (732-8) | 0.12 |
| Lymphocytes/100 leukocytes in Blood (26478-8) | 0.06 |
| Lymphocytes/100 leukocytes in Blood by Automated count (736-9) | 0.10 |
| Lymphocytes/100 leukocytes in Blood by Manual count (737-7) | 0.09 |
| MCH [Entitic mass] (28539-5) | 0.05 |
| MCH [Entitic mass] by Automated count (785-6) | 0.06 |
| MCHC [Mass/volume] (28540-3) | 0.09 |
| MCHC [Mass/volume] by Automated count (786-4) | 0.09 |
| MCV [Entitic volume] (30428-7) | 0.06 |
| MCV [Entitic volume] by Automated count (787-2) | 0.07 |
| Magnesium [Mass/volume] in Serum or Plasma (19123-9) | 0.17 |
| Metamyelocytes [# /volume] in Blood (30433-7) | 0.20 |
| Metamyelocytes/100 leukocytes in Blood (28541-1) | 0.16 |
| Microalbumin [Mass/volume] in Urine (14957-5) | 0.21 |
| Monocytes [# /volume] in Blood (26484-6) | 0.09 |
| Monocytes [# /volume] in Blood by Automated count (742-7) | 0.12 |
| Monocytes [# /volume] in Blood by Manual count (743-5) | 0.13 |
| Monocytes/100 leukocytes in Blood (26485-3) | 0.11 |
| Monocytes/100 leukocytes in Blood by Automated count (5905-5) | 0.12 |
| Monocytes/100 leukocytes in Blood by Manual count (744-3) | 0.22 |
| Myelocytes/100 leukocytes in Blood (26498-6) | 0.23 |
| Natriuretic peptide B [Mass/volume] in Serum or Plasma (30934-4) | 0.08 |
| Natriuretic peptide.B prohormone N-Terminal [Mass/volume] in Serum or Plasma (33762-6) | 0.08 |
| Neutrophils [# /volume] in Blood (26499-4) | 0.08 |
| Neutrophils [# /volume] in Blood by Automated count (751-8) | 0.10 |
| Neutrophils [# /volume] in Blood by Manual count (753-4) | 0.11 |
| Neutrophils.immature/100 leukocytes in Blood (53797-7) | 0.13 |
| Neutrophils/100 leukocytes in Blood (26511-6) | 0.09 |
| Neutrophils/100 leukocytes in Blood by Automated count (770-8) | 0.09 |
| Neutrophils/100 leukocytes in Blood by Manual count (23761-0) | 0.16 |
| Nucleated erythrocytes [# /volume] in Blood (30392-5) | 0.05 |
| Nucleated erythrocytes [# /volume] in Blood by Automated count (771-6) | 0.10 |
| Nucleated erythrocytes [# /volume] in Blood by Manual count (772-4) | 0.06 |
| Nucleated erythrocytes/100 leukocytes [Ratio] in Blood (19048-8) | 0.07 |
| Nucleated erythrocytes/100 leukocytes [Ratio] in Blood by Automated count (58413-6) | 0.06 |
| Nucleated erythrocytes/100 leukocytes [Ratio] in Blood by Manual count (18309-5) | 0.23 |
| Osmolality of Serum or Plasma by calculation (18182-6) | 0.44 |
| Oxygen [Partial pressure] in Arterial blood (2703-7) | 0.26 |
| Oxygen [Partial pressure] in Blood (11556-8) | 0.23 |
| Oxygen saturation Calculated from oxygen partial pressure in Arterial blood (51733-4) | 0.17 |
| Oxygen saturation in Arterial blood (2708-6) | 0.35 |
| Oxygen saturation in Blood (20564-1) | 0.33 |
| Parathyrin.intact [Mass/volume] in Serum or Plasma (2731-8) | 0.30 |
| Percentage change in weight (248347000) | 0.24 |
| Phosphate [Mass/volume] in Serum or Plasma (2777-1) | 0.18 |
| Platelet distribution width [Entitic volume] in Blood by Automated count (32207-3) | 0.01 |
| Platelet mean volume [Entitic volume] in Blood (28542-9) | 0.10 |
| Platelet mean volume [Entitic volume] in Blood by Automated count (32623-1) | 0.11 |
| Platelet mean volume [Entitic volume] in Blood by Rees-Ecker (776-5) | 0.30 |
| Platelets [# /volume] in Blood (26515-7) | 0.15 |
| Platelets [# /volume] in Blood by Automated count (777-3) | 0.14 |
| Platelets [# /volume] in Blood by Manual count (778-1) | 0.24 |
| Platelets reticulated/100 platelets in Blood by Automated count (71693-6) | 0.10 |
| Polymorphonuclear cells/100 leukocytes in Blood (34999-3) | 0.25 |
| Potassium [Moles/volume] in Blood (6298-4) | 0.21 |
| Potassium [Moles/volume] in Serum or Plasma (2823-3) | 0.20 |
| Potassium [Moles/volume] in Serum, Plasma or Blood (77142-8) | 0.12 |
| Prealbumin [Mass/volume] in Serum or Plasma (14338-8) | 0.14 |
| Procalcitonin [Mass/volume] in Serum or Plasma (33959-8) | 0.34 |
| Prolactin [Mass/volume] in Serum or Plasma (2842-3) | 0.18 |
| Promyelocytes [# /volume] in Blood (26523-1) | 0.07 |
| Promyelocytes/100 leukocytes in Blood (26524-9) | 0.05 |
| Prostate Specific Ag Free [Mass/volume] in Body fluid (59239-4) | 0.05 |
| Prostate Specific Ag Free [Mass/volume] in Serum or Plasma (10886-0) | 0.10 |
| Prostate specific Ag [Mass/volume] in Serum or Plasma (2857-1) | 0.12 |
| Prostate specific Ag [Mass/volume] in Serum or Plasma by Detection limit <= 0.01 ng/mL (35741-8) | 0.08 |
| Protein [Mass/time] in 24 hour Urine (2889-4) | 0.08 |
| Protein [Mass/volume] in Serum or Plasma (2885-2) | 0.11 |
| Protein [Mass/volume] in Urine (2888-6) | 0.28 |
| Protein.monoclonal [Mass/volume] in Serum or Plasma by Electrophoresis (33358-3) | 0.15 |
| Protein.monoclonal band 1 [Mass/volume] in Serum or Plasma by Electrophoresis (51435-6) | 0.23 |
| Prothrombin time (PT) (5902-2) | 0.11 |
| Prothrombin time (PT) in Control Platelet poor plasma by Coagulation assay (5901-4) | 0.04 |
| Pulse rate (78564009) | 0.19 |
| Respiratory rate (86290005) | 0.20 |
| Reticulocytes [# /volume] in Blood (14196-0) | 0.12 |
| Reticulocytes/100 erythrocytes in Blood (4679-7) | 0.16 |
| Reticulocytes/100 erythrocytes in Blood by Automated count (17849-1) | 0.22 |
| Rheumatoid factor [Units/volume] in Serum or Plasma (11572-5) | 0.21 |
| Segmented neutrophils [# /volume] in Blood (30451-9) | 0.10 |
| Segmented neutrophils [# /volume] in Blood by Manual count (768-2) | 0.09 |
| Segmented neutrophils/100 leukocytes in Blood (26505-8) | 0.09 |
| Segmented neutrophils/100 leukocytes in Blood by Manual count (769-0) | 0.12 |
| Sodium [Moles/volume] in Blood (2947-0) | 0.15 |
| Sodium [Moles/volume] in Serum or Plasma (2951-2) | 0.14 |
| Sodium [Moles/volume] in Serum, Plasma or Blood (77139-4) | 0.35 |
| Sodium [Moles/volume] in Urine (2955-3) | 0.28 |
| Specific gravity of Urine (2965-2) | 0.39 |
| Specific gravity of Urine by Automated test strip (53326-5) | 0.15 |
| Specific gravity of Urine by Refractometry automated (50562-8) | 0.30 |
| Specific gravity of Urine by Test strip (5811-5) | 0.26 |
| Systolic blood pressure (271649006) | 0.21 |
| Testosterone [Mass/volume] in Serum or Plasma (2986-8) | 0.14 |
| Thyroglobulin Ab [Units/volume] in Serum or Plasma (8098-6) | 0.07 |
| Thyroglobulin [Mass/volume] in Serum or Plasma (3013-0) | 0.13 |
| Thyroid hormone uptake (T-uptake) in Serum or Plasma (74793-1) | 0.25 |
| Thyrotropin [Units/volume] in Serum or Plasma (3016-3) | 0.19 |
| Thyrotropin [Units/volume] in Serum or Plasma by Detection limit <= 0.005 mIU/L (11580-8) | 0.17 |
| Thyrotropin [Units/volume] in Serum or Plasma by Detection limit <= 0.05 mIU/L (11579-0) | 0.05 |
| Thyroxine (T4) [Mass/volume] in Serum or Plasma (3026-2) | 0.21 |
| Thyroxine (T4) free [Mass/volume] in Serum or Plasma (3024-7) | 0.16 |
| Thyroxine (T4) free index in Serum or Plasma by calculation (32215-6) | 0.32 |
| Transferrin [Mass/volume] in Serum or Plasma (3034-6) | 0.14 |
| Triglyceride [Mass/volume] in Serum or Plasma (2571-8) | 0.16 |
| Triiodothyronine (T3) Free [Mass/volume] in Serum or Plasma (3051-0) | 0.19 |
| Triiodothyronine (T3) [Mass/volume] in Serum or Plasma (3053-6) | 0.21 |
| Troponin I.cardiac [Mass/volume] in Blood (42757-5) | 0.27 |
| Troponin I.cardiac [Mass/volume] in Serum or Plasma (10839-9) | 0.07 |
| Troponin T.cardiac [Mass/volume] in Serum or Plasma (6598-7) | 0.16 |
| Urate [Mass/volume] in Serum or Plasma (3084-1) | 0.16 |
| Urea nitrogen [Mass/volume] in Blood (6299-2) | 0.12 |
| Urea nitrogen [Mass/volume] in Serum or Plasma (3094-0) | 0.12 |
| Urea nitrogen/Creatinine [Mass Ratio] in Blood (44734-2) | 0.25 |
| Urea nitrogen/Creatinine [Mass Ratio] in Serum or Plasma (3097-3) | 0.12 |
| Urobilinogen [Mass/volume] in Urine (3107-0) | 0.11 |
| Urobilinogen [Mass/volume] in Urine by Test strip (20405-7) | 0.12 |
| Urobilinogen [Units/volume] in Urine by Test strip (19161-9) | 0.22 |
| Vancomycin [Mass/volume] in Serum or Plasma --trough (4092-3) | 0.27 |
| Variant lymphocytes [# /volume] in Blood (26477-0) | 0.25 |
| aPTT in Platelet poor plasma by Coagulation assay (14979-9) | 0.12 |
| pH of Arterial blood (2744-1) | 0.34 |
| pH of Urine (2756-5) | 0.25 |
| pH of Urine by Automated test strip (50560-2) | 0.10 |
| pH of Urine by Test strip (5803-2) | 0.21 |

**Supplementary Table 2.** LIFE’s performance across cancer subtypes in terms of mean absolute error. The performance of LIFE was relatively unaffected by the tumor’s origin or stage, exhibiting strong performance even in cases of relatively benign diseases such as early-stage breast cancer. These results suggest that LIFE should generalize to patients without cancer as well. The average performance was assessed based on a selection of 25 routinely conducted laboratory tests, following the same methodology as outlined in the main manuscript for Table 1. The 4 cancer types shown in the table were selected based on the number of samples in the dataset, while all the rest were categorized as “others”. All data pertaining to the tumor’s origin and stage were curated from clinical notes by professional abstractors.

|  |  | Mean Absolute Error |  |  |  |
| --- | --- | --- | --- | --- | --- |
|  |  | Stage 1 | Stage 2 | Stage 3 | Stage 4 |
| Tumor's Origin | Breast | 0.14 | 0.14 | 0.14 | 0.15 |
|  | Colorectal | 0.15 | 0.14 | 0.14 | 0.15 |
|  | Head and Neck | 0.17 | 0.14 | 0.16 | 0.16 |
|  | Lung | 0.15 | 0.14 | 0.16 | 0.16 |
|  | Others | 0.15 | 0.14 | 0.14 | 0.15 |

